# Investigative needle core biopsies for multi-omics in Glioblastoma

**DOI:** 10.1101/2023.12.29.23300541

**Authors:** Kenny K.H. Yu, Sreyashi Basu, Gerard Baquer, Ryuhjin Ahn, Jennifer Gantchev, Sonali Jindal, Michael S. Regan, Zaki Abou-Mrad, Michael C. Prabhu, Marc J. Williams, Alicia D. D’Souza, Seth W. Malinowski, Kelsey Hopland, Yuval Elhanati, Sylwia A. Stopka, Alexei Stortchevoi, Zhong He, Jingjing Sun, Yulong Chen, Alexsandra B. Espejo, Kin Hoe Chow, Smitha Yerrum, Pei-Lun Kao, Brittany Parker Kerrigan, Lisa Norberg, Douglas Nielsen, The GBM TeamLab, Vinay K. Puduvalli, Jason Huse, Rameen Beroukhim, Yon Son Betty Kim, Sangeeta Goswami, Adrienne Boire, Sarah Frisken, Michael J. Cima, Matthias Holdhoff, Calixto-Hope G. Lucas, Chetan Bettegowda, Stuart S. Levine, Tejus A. Bale, Cameron Brennan, David A. Reardon, Frederick F. Lang, E. Antonio Chiocca, Keith L. Ligon, Forest M. White, Padmanee Sharma, Viviane Tabar, Nathalie Y. R. Agar

## Abstract

Glioblastoma (GBM) is a primary brain cancer with an abysmal prognosis and few effective therapies. The ability to investigate the tumor microenvironment before and during treatment would greatly enhance both understanding of disease response and progression, as well as the delivery and impact of therapeutics. Stereotactic biopsies are a routine surgical procedure performed primarily for diagnostic histopathologic purposes. The role of investigative biopsies – tissue sampling for the purpose of understanding tumor microenvironmental responses to treatment using integrated multi-modal molecular analyses (‘Multi-omics”) has yet to be defined. Secondly, it is unknown whether comparatively small tissue samples from brain biopsies can yield sufficient information with such methods. Here we adapt stereotactic needle core biopsy tissue in two separate patients. In the first patient with recurrent GBM we performed highly resolved multi-omics analysis methods including single cell RNA sequencing, spatial-transcriptomics, metabolomics, proteomics, phosphoproteomics, T-cell clonotype analysis, and MHC Class I immunopeptidomics from biopsy tissue that was obtained from a single procedure. In a second patient we analyzed multi-regional core biopsies to decipher spatial and genomic variance. We also investigated the utility of stereotactic biopsies as a method for generating patient derived xenograft models in a separate patient cohort. Dataset integration across modalities showed good correspondence between spatial modalities, highlighted immune cell associated metabolic pathways and revealed poor correlation between RNA expression and the tumor MHC Class I immunopeptidome. In conclusion, stereotactic needle biopsy cores are of sufficient quality to generate multi-omics data, provide data rich insight into a patient’s disease process and tumor immune microenvironment and can be of value in evaluating treatment responses.

**One sentence summary:** Integrative multi-omics analysis of stereotactic needle core biopsies in glioblastoma

## Introduction

Glioblastoma (GBM) is an aggressive primary brain cancer with limited treatment options (*1*). Despite numerous clinical trials to date, the standard of care remains maximal surgical resection followed by combination chemotherapy and radiotherapy (*2–4*). Apart from tumor-treating fields (*4*), few novel therapies have been approved. Unfortunately, despite aggressive multimodality treatments, the prognosis remains poor. While immunotherapies in other solid cancers have shown significant improvements in overall survival, trials in GBM which include targeted and immunotherapies have been met with limited success (*5, 6*).

Standard clinical endpoints such as overall survival, progression-free survival or radiological evidence of progression are coarse markers of treatment response, and there is a strong need to better understand how the tumor, the host immune system and the tumor microenvironment respond to therapies. There are currently few opportunities to obtain additional tissue samples to understand tumor response during treatment other than at initial surgery in the form of either biopsy or resection. This lack of visibility into tumor and immune co-evolution under treatment is a fundamental roadblock to our ability to develop and advance new GBM therapies.

Stereotactic biopsies are a routine neurosurgical procedure typically only performed for diagnostic purposes to obtain a tissue diagnosis through basic histological and molecular evaluation (*7, 8*). The trend however towards obtaining additional molecular data besides traditional hematoxylin and eosin (H&E), and immunohistochemical (IHC) staining has begun in earnest and many academic medical centers have incorporated gene panel sequencing such as MSK-IMPACT (*9*), DFCI Oncopanel or other solid tumor panels (*10, 11*) from needle core biopsies as part of the routine clinical diagnostic workflow. The amount of clinical material obtained from these core biopsies is small (<50mg). The uni-directional cutting face of the biopsy needle however, permits multiple cores to be taken from the same site from different directions by rotating along its axis (Figure 1A).

**Fig. 1.**
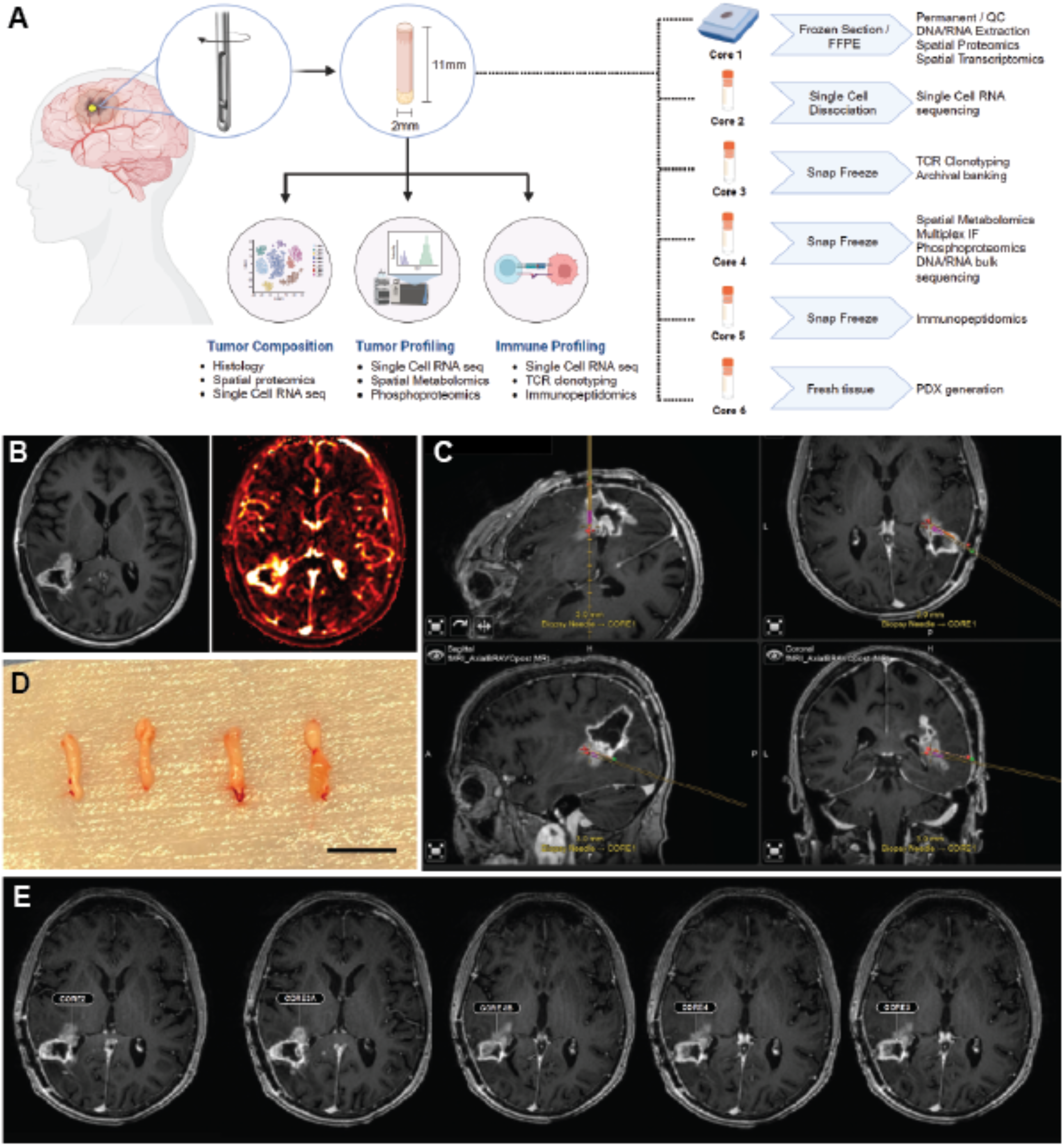
Intra-operative collection and coordination of sample cores distribution and analyses. A) Schematic diagram demonstrating multiple core sampling and proposed analysis methods. Note that the biopsy needle can rotate along its axis and perform multiple biopsies in from the same fixed position. B) MR Imaging of patient undergoing resection of lesion. Left, T1 weighted imaging with contrast. Right, MR perfusion imaging demonstrating “hotspot” on the anteromedial portion of tumor, destined for resection. C) Intra-operative neuro navigated imaging demonstrating the trajectory and position of biopsy needle in 3 orthogonal planes and in-line view. D) Representative pictures of multiple biopsy cores (scale bar = 10mm). E) Core sampling locations geospatially mapped back to original MR imaging using 3D slicer software.

The role of investigative biopsies – i.e., biopsies where the primary goal is to collect systematic, detailed multi-modal tumor related information for the purposes of evaluating treatment response, has been under-explored (*12*). Development of GBM immunotherapies will require information on the spectrum and profile of intra-tumoral immune cells present, how these populations shift in size and what immune cell activation states are present during treatment. This information may have important implications for decisions regarding treatment response, disease progression and prognosis. The ability to track tumor and stromal populations and resolve cell-cell signaling pathways via spatial and single cell technologies could potentially offer a comprehensive overview of the tumor fitness landscape under different therapeutic modalities (*13*).

The application of multi-omics technologies to the GBM tumor microenvironment (TME) represents a powerful and unbiased approach to generate unique insight into tumor-stromal interactions and tumor organizational structure (*14*); single cell RNA sequencing combined with spatial metabolomics analysis has shown that metabolites in the TME can shape the tumor immune environment, through purinergic signaling pathways, involving ATP and adenosine in tumor-microglial interaction via CD73 and CD39 respectively (*15*) or direct immunosuppressive effects via oncometabolites on T-cells (*16*); phosphoproteomics and immunopeptidomics techniques have shown how the signaling pathways and the antigenic peptide repertoire can alter under treatment (*17–20*) thus shaping the systemic immune response to tumor. We selected 6 assays as part of our multi-modal analysis panel: 1) scRNA sequencing, 2) spatial transcriptomics, 3) spatial proteomics, 4) spatial metabolomics, 5) phosphoproteomics and 6) HLA Class I immunopeptidomics. Application of these selected multi-omics technologies however to the analysis of needle core biopsy samples poses significant challenges such as the adaptation of assays to accommodate small input volumes, and the primary goal was to demonstrate safety and utility in performing multiple biopsies on a single patient. Given our understanding of tumor heterogeneity in GBM, a secondary objective was to establish methods to measure intra- and inter-regional variance from biopsies within the tumor.

Here we demonstrate the feasibility of this multi-omics approach on needle core biopsies from an individual patient. We developed an optimized workflow to obtain information regarding cellular composition, tissue architecture, cellular transcriptomes, the MHC Class I immunopeptidome and protein activation pathways from needle core biopsies collected during surgery. Secondary integration of multi-modal measurements then allowed us to cross-validate findings and make inferences about immune cell metabolic activity within the TME. The overall clinical and scientific impetus to develop multi-modal analysis methods is driven by the need to understand disease progression and tumor-immune evolution over the course of treatment. The establishment of protocols that can facilitate this has important implications for clinical trials and disease monitoring going forward.

## Results

### Intra-operative spatially localized needle core biopsy sampling

Multiple biopsies were taken from a patient undergoing craniotomy for resection of a recurrent GBM after obtaining informed consent **(Figure 1B)**. The patient had presented with tumor recurrence after completing standard of care first line treatment (**Supplementary Table 1**). After re-opening of the craniotomy and exposure of the target resection area, we obtained sequential biopsies using a biopsy needle registered to a frameless stereotaxy system (BrainLab AG, Germany) in a region of tumor destined for resection, along a single trajectory. We recorded the 3D coordinates using the intra-operative neuronavigation software at each depth and biopsy location (iPlan 3.0 software, BrainLab AG, Germany). Samples were obtained from 4 quadrants where feasible before moving to a separate depth and obtaining additional samples. Five core needle biopsy samples (out of a total of seven cores obtained) were deemed of sufficient quality for downstream analysis **(Figure 1C-E)**. Sample volumes obtained from biopsy needle cores were in the range of 41-44mg. Tissue cores were immediately placed on labelled saline-soaked non-adherent patties (Telfa®, American Surgical Company, USA). All cores were inspected visually to ensure the specimen was solid tissue and not blood clot. The two cores which had the most viable appearance were placed in tissue storage solution (MACS storage buffer, Miltenyi Biotec, Germany) and placed on ice for fresh processing (Core 2). The other cores were assigned according to the flowchart **(Figure 1A)** and were either fixed in formalin (Core 1), or snap frozen using liquid nitrogen vapor (Cores 3-5). Cores were then transported on an expedited basis to partner institutions for processing and analysis. In addition, excess tissue from Core 1 (formalin fixed) was preserved for bulk T-cell receptor clonal analysis **(Supplementary Figure 1)**, excess frozen tissue from Core 4 (frozen) was used to perform bulk RNA sequencing.

### Single cell RNA sequencing of GBM biopsy cores

Single cell RNA sequencing (scRNA) technology permits high resolution gene expression analysis of tissue samples, and has been published extensively in GBM showing glioma cell states (*21–23*), immune cell composition and oncogenic developmental programs (*24–26*). Nearly all studies utilized surgical resection samples, and it is unknown whether current tissue processing protocols for resection samples are suitable for needle biopsy cores (*27*). We therefore adapted our existing protocols to low input volumes. Similar challenges were faced in diffuse midline gliomas such as intrinsic pontine glioma (DIPG), where tissue samples are necessarily limited due to the extreme sensitivity of the brain structures involved. Although scRNAseq was shown to be feasible with low tissue input volumes (*28, 29*), the number of cells sequenced was low (2458 cells from 6 patients)(*30*). More recently multi-location biopsy sampling was combined with scRNAseq (*31*), and in this study, the investigators sequenced 6148 cells from 73 biopsy sites in 13 patients. Overall, the combined sequenced cell yield per biopsy in these studies was in the region of 80-400 cells per biopsy. We estimated however that we would need to aim for 10,000 cells to adequately detect tumor, immune and stromal populations which would require a minimum starting quantity of 50,000 viable cells. Single cell dissociation of brain tumor tissue typically involves mechanical dissociation followed by enzymatic digestion, optionally followed by density gradient separation or myelin removal and red cell lysis (*25*). This process can be standardized through automated tissue trituration and dissociation methods (GentleMACS system, Miltenyi Biotec, Germany). Automated tissue processing has the advantage of providing consistent conditions, however the instrument settings are not optimized for low volume samples. We therefore first performed a direct comparison between manual dissociation versus automated dissociation and showed that manual dissociation yielded approximately ten-fold as many live cells as the automated method, with the average yield per core of approximately 610,000 live cells after manual dissociation compared to 74,000 with automated methods (**Supplementary Table 2**).

A recent study by Marsh et al. suggested that enzymatic dissociation is associated with an ex-vivo activation signature (ExAM) which was absent in mechanical dissociation and is abrogated using protein translational inhibitors (*32*). The original study was performed in normal mouse brains, therefore we sought to determine if this held true for human tumor tissue. To address this, we first performed single cell dissociation and scRNAseq with and without translational inhibitors. We then repeated the experiment with and without enzymatic digestion, with and without inhibitors (**Figure 2A-B, F-G**). Corroborating the report by Marsh et al., translational inhibitors clearly decreased the ExAM signature in the enzymatically dissociated samples, particularly in the myeloid population. Comparison of enzyme versus non-enzymatic mechanical conditions showed the ExAM signature was higher in the mechanically dissociated sample compared to the enzymatic condition. This observation suggests that non-enzymatic effects also contributed to the ExAM signature in the context of GBM tissue. Pure mechanical dissociation alone without enzymes also yielded a much lower number of viable cells, severely hindering the quality of the downstream analysis.

**Fig. 2.**
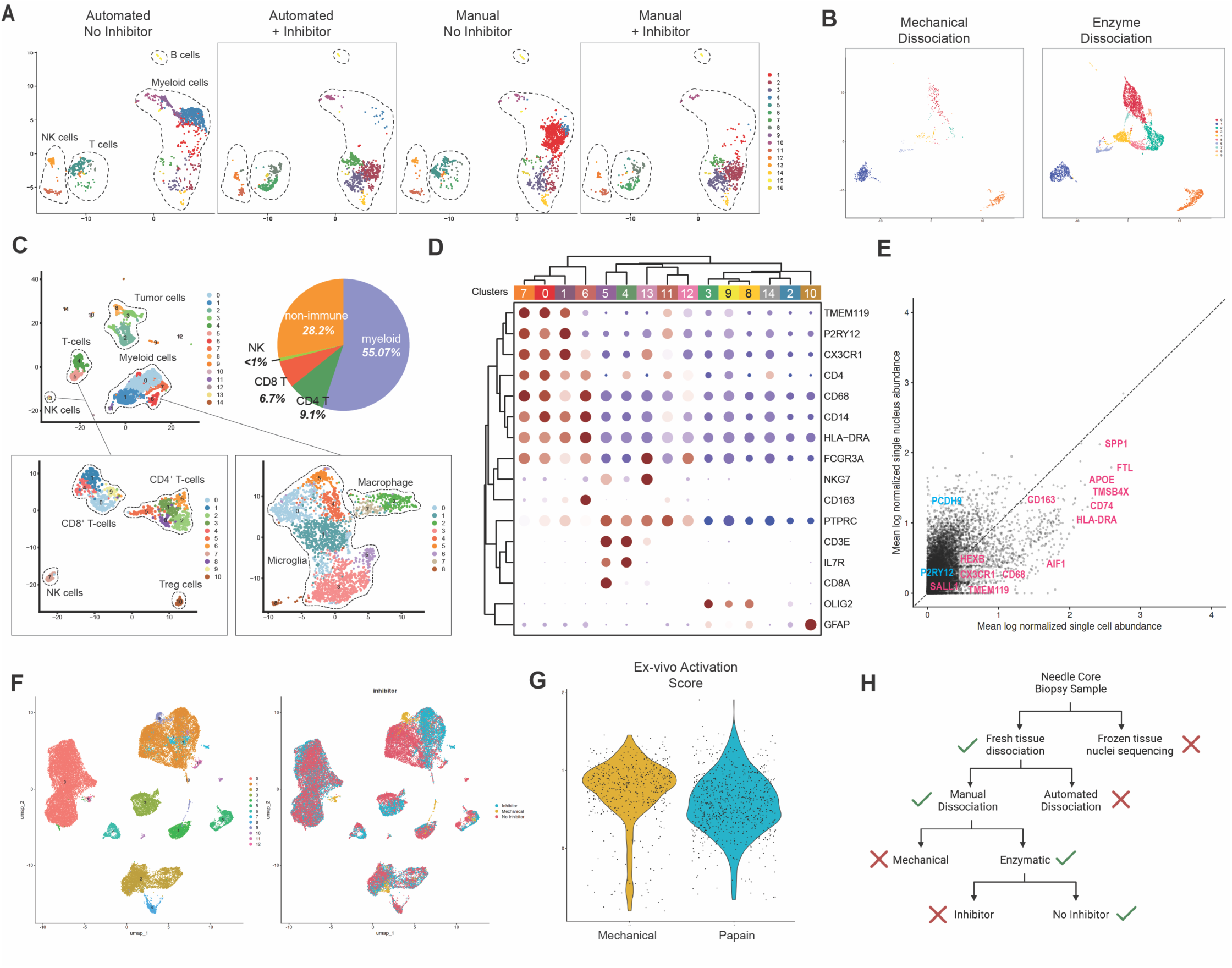
Optimization of single cell workflow for fresh GBM biopsy cores. A) UMAP representation of scRNA data from needle core biopsy samples. B) proportion of cell types. C) Representative markers for identified cell types. D) viable cell yield from needle core biopsy dissociation comparing automated dissociation methods (GentleMACS) versus manual dissociation. E) comparison between frozen tissue derived single cell nuc-seq versus fresh cell single cell sequencing from the same patient. Populations represent a direct comparison between glioma associated macrophages from the same sample. F) Ex-vivo Activation Signature (ExAM) score in sequenced cell populations between inhibitor and no inhibitor conditions. G) Comparison of mechanical (non-enzymatic) and enzymatic dissociation with and without inhibitor. Bottom panel: ExAM score between mechanical versus enzymatic dissociation. H) Decision tree for single cell processing workflow for stereotactic biopsy samples.

Another technical and logistical consideration was to determine whether scRNA should be performed using freshly dissociated single cells or frozen sample derived single nuclei, with the rationale being that frozen samples are easier to transport, have a better chance of preserving nuclei in sufficient numbers for downstream single nuclei RNA sequencing (snRNA), and capture dissociation-susceptible cell types such as neurons. Previous studies comparing these two methods have shown loss of specific gene expression, particularly in microglial cells (*33*). Given that one of our key study objectives was to study the immune myeloid cell composition, we performed pilot experiments in a separate patient with IDH wildtype GBM and compared the yield of CD45 positive population in both conditions which corroborated the findings by Thrupp et al. We found important differences in gene expression between scRNA and snRNA, with key genes associated with glioma associated myeloid cells (GAM) better represented in the scRNA data compared to snRNA (**Figure 2E**). These genes included *CD163*, *CX3CR1*, *CD163*, *AIF1*, *CD68*, *P2RY12* and *SORL1*. In view of our interest in tumor microenvironmental changes to treatment, we therefore concluded that enzymatic fresh tissue dissociation without the use of translational inhibitors for scRNA would be preferable (**Figure 2H**).

### Spatial proteomic profiling of biopsy cores

GBM is a histomorphologically heterogeneous tumor with regions of cellularity, infiltration, microvascular proliferation and necrosis (*34*). Spatial tissue profiling technologies have emerged as a method of identifying intratumoral cellular neighborhoods to gain phenotypic understanding of niche-specific molecular heterogeneity of GBM (*35*). We performed highly multiplexed microscopy using Co-detection by indexing (CODEX, Akoya Biosciences, USA). A panel of 51 antibodies was validated for tumor microenvironment specific protein markers on FFPE tissue to identify immune cell types, tumor, vascular and stromal markers along with multiple functional markers (**Figure 3A**, **Table 1**). The distance between of specific immune cells from GFAP+ cells was evaluated by near neighbor analysis for the CD3+ T cells and CD68+ macrophages. Macrophages were more abundant and closer to the GFAP+ cells suggesting closer interaction between two cell types when compared to T-cells (**Figure 3B**, **table 1**). Clustering of different cellular subpopulations showed an abundance of GFAP+ cells (33%) closely followed by myeloid cells (18%). Previous studies have shown that lymphocytic infiltration tends to be limited to the perivascular spaces, however in our sample there was a paucity of T cells and comprised of only ∼1% of all cell type population (**Figure 3C**) with no clear relationship to the vasculature. Further, a total of 15 cellular subtypes were identified (**Supplementary Figure 2**). These cell clusters were further interrogated to characterize cellular neighborhoods (CN) that were the re-mapped to the original image to reveal the spatial organization of CN (**Figure 3D-E**). A total of six different CN were identified with tumor cells (CN1) being most frequent and surrounded by proliferative tumor and myeloid cell rich neighborhood (CN2). Additional CNs including vasculature associated immune cells (CN3), macrophage and microglial cells (CN4), myeloid and B cells (CN5), and GFAP+ cells in association with reactive astrocytes (CN6) were also observed. Defining spatial organization can provide insight into interactions of the different cell subtypes within the tumor ecosystem and the potential impact of drug treatment on tumor evolution and cell-cell cross communication.

**Fig. 3.**
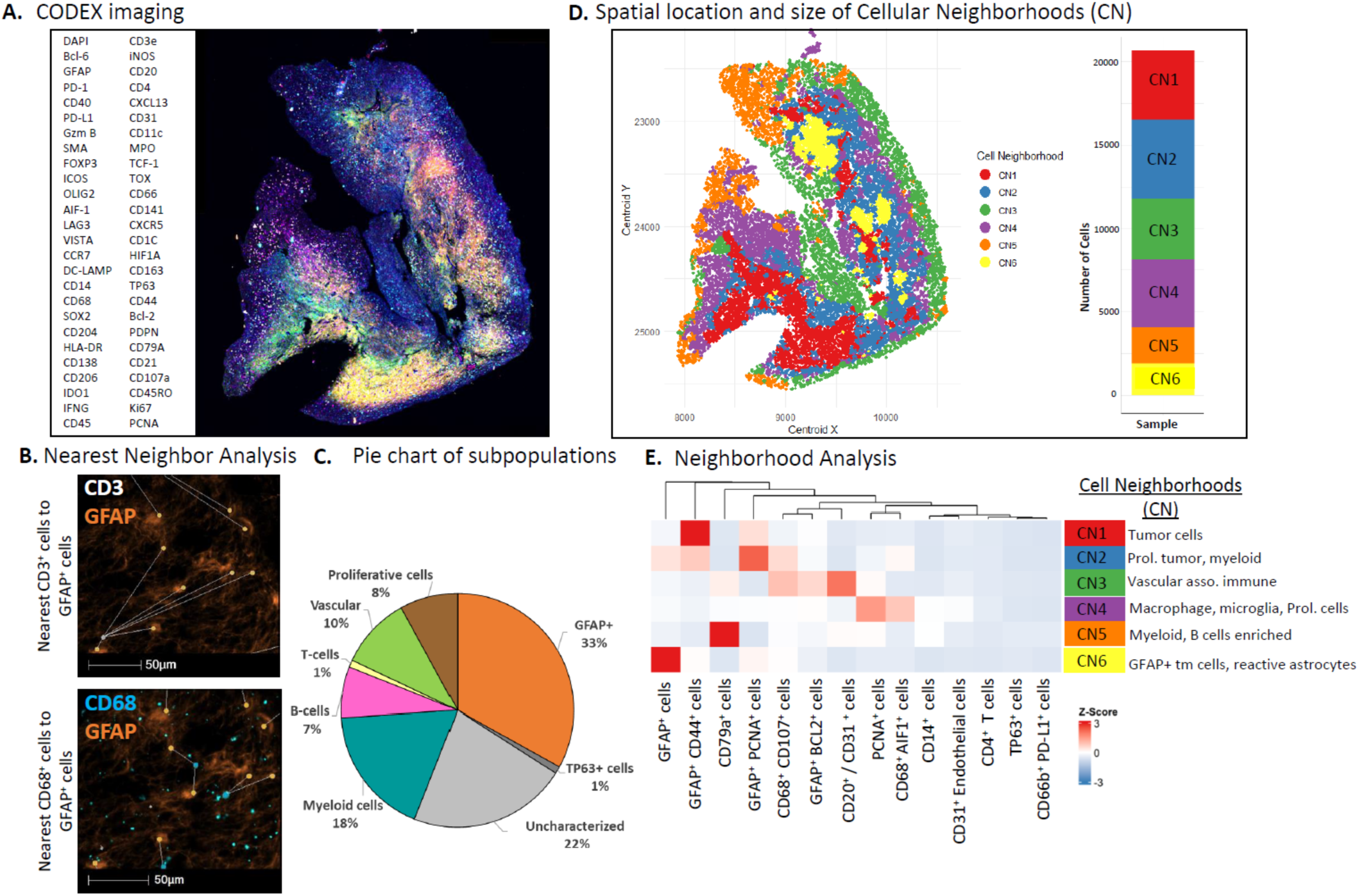
A) CODEX panel and representative image showing all biomarker staining. B) Nearest Neighbor analysis showing distance of CD3+ T cells and CD68+ macrophages to the GFAP+ cells. C) Pie chart showing different subpopulations present within the pilot tissue sample. D) Spatial location of Cellular Neighborhood (CN) as seen on the biopsy tissue. Stack plot represents frequency of different CN. E) Neighborhood analysis show different cellular clusters contributing to form different CN.

### Cell signaling with proteomics and phosphoproteomics

Protein tyrosine phosphorylation (pTyr) can regulate protein activity, interactions, localization, and stability. Signaling networks mediated by pTyr control a variety of cellular processes in response to intracellular and extracellular (e.g., environmental) perturbations. Aberrant pTyr signaling is known to be oncogenic, with many tyrosine kinases identified as potent oncogenes, and conversely protein tyrosine phosphatases are known to be tumor suppressors. Moreover, tyrosine phosphorylation regulates adaptive and innate immune cell activity through cell surface receptors, JAK/STAT signaling, and immune checkpoint proteins such as SIRPa and the Siglecs. Receptor Tyrosine Kinase (RTK) pathways such as Epidermal Growth Factor (EGF) and Platelet Derived Growth Factor (PDGF) alpha and beta family of associated receptors are frequently aberrantly activated in GBM and result in an oncogenic phenotype (*36, 37*). STAT3 signaling has also been implicated in GAM polarization in GBM and recruitment of regulatory T-cells (*38, 39*). We utilized mass spectrometry-based phosphotyrosine phosphoproteomics to identify tyrosine phosphorylated proteins in the patient tumor and capture the tumor cell-instrinsic and immune signaling components. Several 10-micron thick sections from Core 5 were processed to yield tryptic peptides which were subsequently enriched for pTyr-containing peptides by immunoprecipitation and phosphopeptide enrichment. Enriched pTyr peptides were then analyzed by liquid chromatography-tandem mass spectrometry (LC-MS/MS) on an Exploris 480 Orbitrap mass spectrometer. This analysis yielded 256 unique phosphorylation sites (171 pTyr and 85 pSer/pThr) on proteins ranging from receptor tyrosine kinases such as EGFR and their downstream signaling networks, including the ERK mitogen-activated protein kinases (MAPKs) to immune cell-specific proteins including CD226 (Y322), CD84 (Y279, Y296, Y316), and Siglec-9 (Y456)(**Figure 4A**, **Supplementary Table 3**). The area-under-the-curve (AUC) was quantified for the chromatographic elution profile for each phosphopeptide to evaluate the respective abundance of the various phosphorylation signals. This analysis highlighted the most abundant signals, including activation loop phosphorylation sites on well-known oncogenes such as the ERK MAPKs and Src-family kinases (SFKs) (Lyn/Hck and Yes/Src/Fyn/Lck) as well as activating sites on p38 MAPK and the STAT3 transcription factor, which has been implicated in driving tumor progression in GBM. Although EGFR phosphorylation was detected, only a single site (Y1173) was identified at relatively low abundance compared to SFKs or ERK MAPKs, suggesting that this tumor was more likely to be driven by SFKs than EGFR signaling. In addition to the oncogenic signals, a large number of pTyr sites are indicative of innate immune infiltration into the tumor, including sites on the immunoglobulin epsilon receptor (FcER) and immunoglobulin gamma receptor (FcGR), as well as sites on the Tyro receptor binding protein (TYROBP), Siglec-9 and PECAM1. Supernatant from pTyr-immunoprecipitation was fractionated using a high pH gradient for global phosphoproteomic (1519 pSer/pThr) and proteomic (2746 unique proteins) analysis (**Figure 4A**, **Supplementary Table 3**). The quantified proteins and phosphoproteins are involved in a plethora of biological pathways (e.g., apoptosis, translation, neuron-projection development, metabolism, etc.) that cover various hallmarks of cancer (**Figure 4B**). The phosphorylation status of critical immune checkpoint proteins such as PD-L1 (S283) and VISTA (S235) showed that these receptors were present and engaged in the tissue. Additionally, 68 proteins and 38 phosphorylation sites implicated in several metabolic pathways (e.g., pyruvate, ATP, NADP, and fatty acid metabolic pathways) were quantified in the biopsied sample. Taken together, we demonstrated that comprehensive phosphoproteomic and proteomic analysis could be done on a small amount of biopsy to shed light on the complex signaling networks formed by tumor and the surrounding microenvironment (**Figure 4B**).

**Fig. 4.**
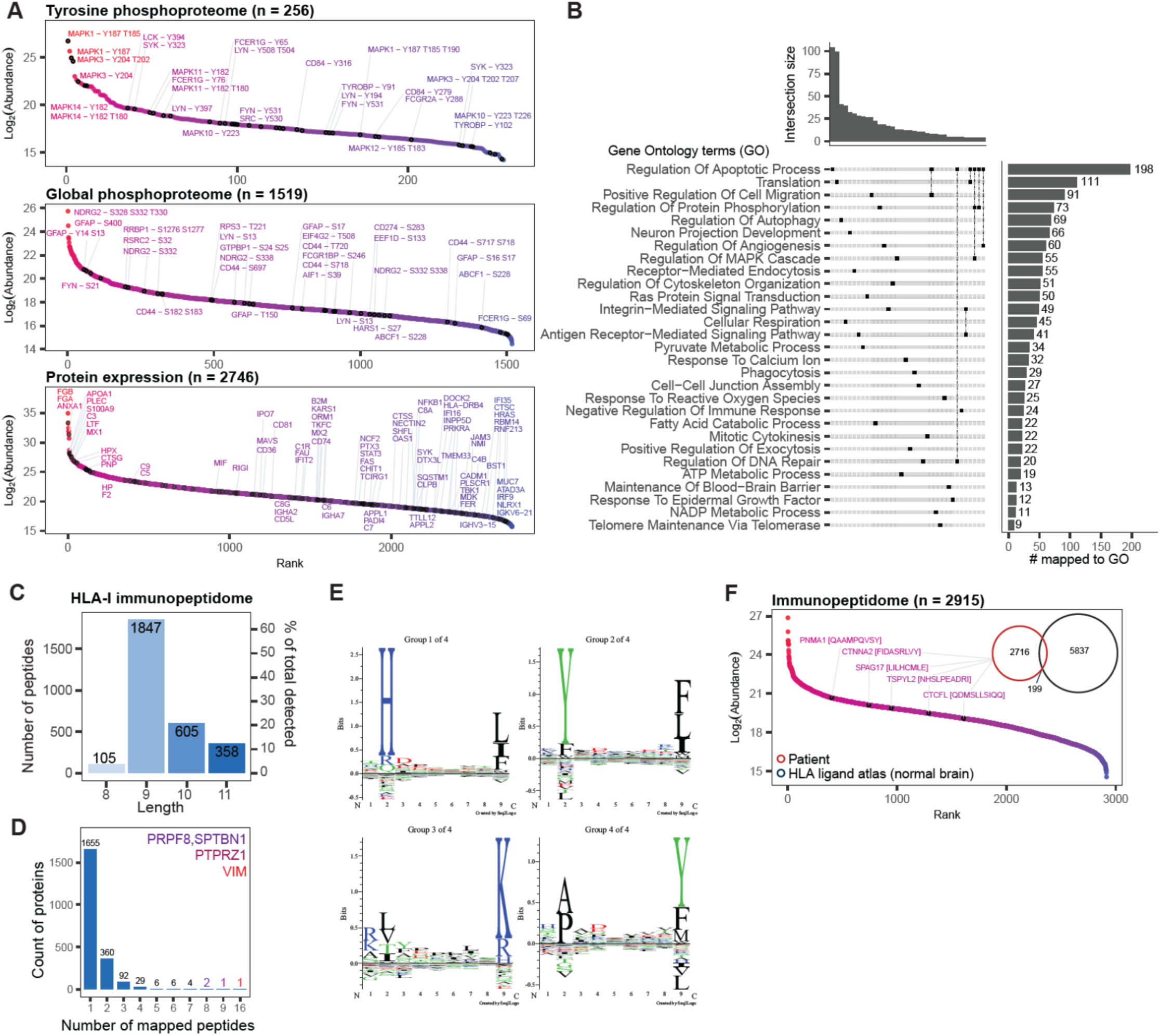
The depth of proteomic, phosphoproteomic, and immunopeptidomic analysis on GBM core biopsy. A) Unique proteins and phosphoproteins quantified are ranked by their abundance. The names of select proteins and phosphoproteins associated with tumor progression and immune infiltration are shown. B) Gene ontology terms associated with proteins and phosphoproteins that were quantified. Groups that had intersection sizes above 3 are shown. C) The number of 8 to 11-mer peptides identified by immunopeptidomic analysis. D) The number of proteins that gave rise to MHC peptides. E) Motif analysis of the quantified immunopeptidome using Gibbs clustering. F) MHC peptides quantified from the core biopsy were compared to the HLA Ligand Atlas of the benign human brain. Previously reported tumor-associated antigens are labeled along the immunopeptides that were ranked based on their abundance.

### Immunopeptidomics on GBM biopsy cores

Immunological control of tumor growth is based upon the recognition of MHC Class I and Class II peptides by CD8+ and CD4+ T cells respectively. Immune checkpoint inhibitors have been unsuccessful in GBM clinical trials. Results from targeted immunotherapies with MHC Class I neoantigens or tumor-associated antigens have, however, suggested potential therapeutic benefit in selected patients, especially if the treatment can be matched to the immunopeptide expression profile for a given patient (*40*). To determine the MHC Class I profile for the current patient, a frozen core (Core 5) was subjected to mass spectrometry-based immunopeptidomic analysis. Following homogenization and cell lysis which yielded 1.95 mg of lysate, MHC class I peptide complexes were isolated using W6/32 antibody. MHC I peptides were then released by low pH and enriched by molecular weight cut-off filter prior to the analysis by LC-MS/MS. This analysis led to the identification of 2,915 MHC class I peptides, of which 1,847 were 9-mers and 605 were 10-mers (**Figure 4C**). Additionally, VIM, PTPRZ1, PRPF8, and SPTBN1 were highly represented in the immunopeptidome, contributing 16, 9, 8, and 8 epitopes, respectively, suggesting their high expression in the biopsy core (**Figure 4D**). These proteins have been previously implicated in glioblastoma invasion and progression (*41–43*). Gibbs clustering of the data highlighted four MHC peptide motifs (**Figure 4E**). Of the 2,915 peptides detected, 199 were found within the HLA Ligand Atlas, a comprehensive collection of HLA ligands presented on benign tissues, suggesting that these peptides may represent those that are enriched in malignant tissues (*44*). In line with this, previously characterized tumor-associated antigens were identified in the patient-specific immunopeptidome, again highlighting the ability of this technique to detect antigens presented in glioblastoma (**Figure 4F**).

### Spatial Metabolite Imaging

The integration of H&E, t-CyCIF and MALDI-MSI data of consecutive biopsy tissue sections reveals insights into the spatial context of immune cell metabolism in GBM (**Figure 5**). Adenosine triphosphate (ATP), an essential compound in metabolism and pathway signaling (*45, 46*) is generated by aerobic glycolysis and the tricarboxylic acid (TCA) cycle in GBM (*47*). Our analysis demonstrates ATP positive pixels acquired with MALDI-MSI in CD45-regions shown in t-CyCIF analysis (**Figure 5A**). The extracellular breakdown of ATP into adenosine, called purinergic signaling, plays a role in pathogenesis and enhanced chemoresistance in GBM (*48*). High CD73 expression, the ectonuclease responsible for the conversion of AMP to adenosine, is found on CD45+ immune cells that suppress immune activity (*49*) and results in decreased levels of ATP. Conversely, the catabolism of ATP is exceedingly slow in glioma cells, resulting in accumulation of ATP in proximity to tumor cells (*50*). On the other hand, malate and linoleic acid metabolism are enriched in regions of CD45+. Although the exact mechanisms in GBM are unclear, high levels of either malate or linoleic acid lead to metabolic reprogramming in the mitochondria (*51, 52*). Linoleic acid, a highly consumed polyunsaturated fatty acid, has been shown to improve antitumor function of CD8+ T cells through metabolic reprogramming and facilitates superior cytotoxic function (*53*). Integration of spatial proteomics and spatial metabolomics show enriched metabolites in CD45+ and CD45-regions (**Figure 5B**) that correspond to metabolic pathways throughout the biopsy sample (**Figure 5C, D**). CD45-regions are enriched for ubiquinone and other terpenoid-quinone biosynthesis; caffeine metabolism; arginine and proline metabolism; aminoacyl-tRNA biosynthesis; tryptophan and tyrosine metabolism. GBMs rely on glucose for metabolic fuel. They also, however utilize amino acids such as glutamine and glutamate (*54*). Reprogrammed glutamine is essential for proliferation and survival of cancer cells which also yields an increase in amino acids, TCA cycle intermediates and fatty acid biosynthesis (*55*). Our results consistently demonstrate enriched amino acid pathways in the CD45+ regions such as taurine and hypotaurine metabolism, valine, leucine and isoleucine biosynthesis, arginine biosynthesis, linoleic metabolism and D-glutamine and D-glutamate metabolism.

**Fig. 5.**
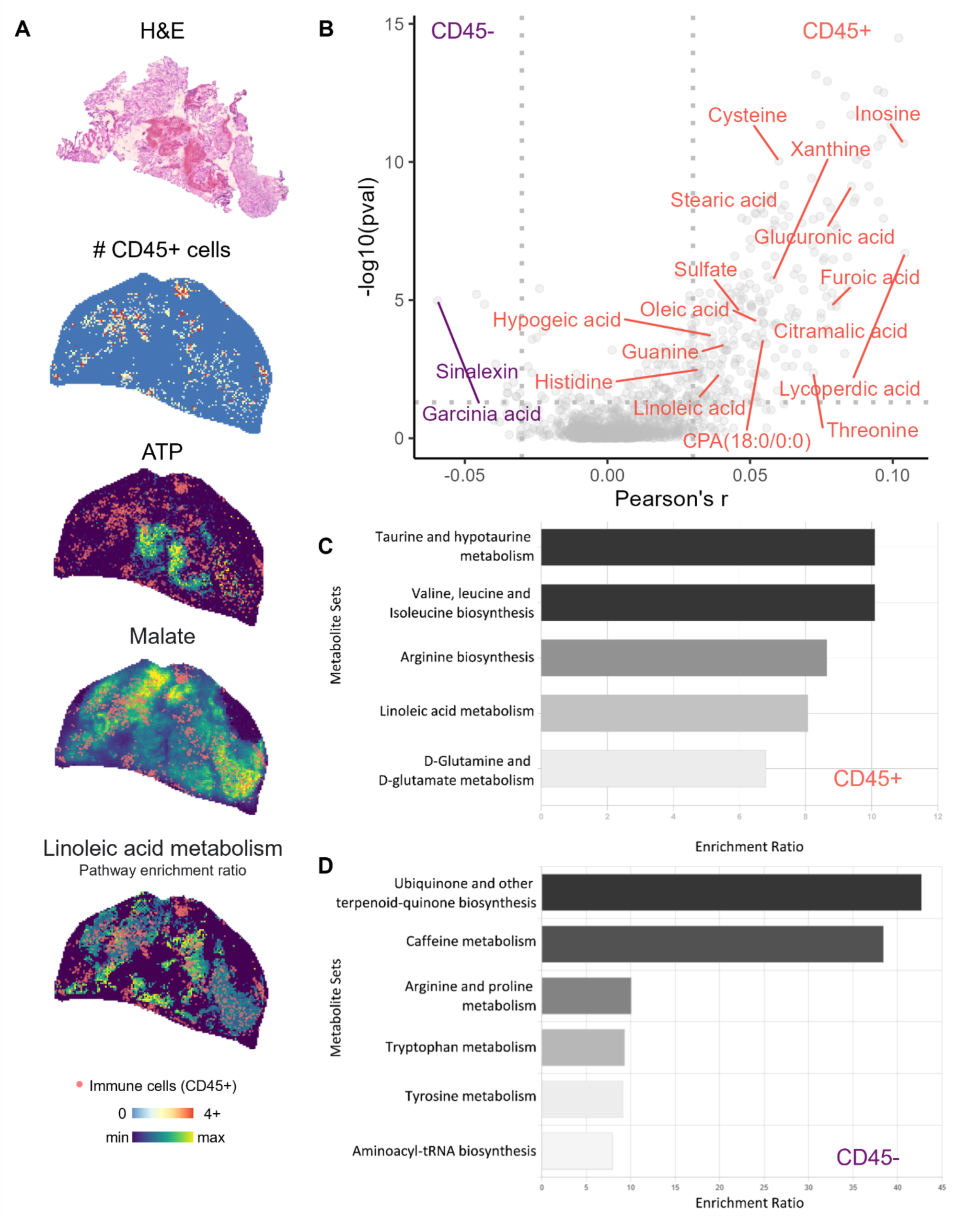
Integrated spatial proteomics and metabolomics on GBM biopsy core #4. A) Hematoxylin and Eosin staining, and corresponding spatial distribution of immune cells (CD45+), Adenosine triphosphate (ATP), Malate, and Linoleic acid metabolism enrichment with immune cells (CD45+) overlayed. The distribution of ATP is anticorrelated, while Malate and Linoleic acid metabolism show strong spatial colocalization to immune cells. B) Comparison of metabolism between immune and non-immune regions of the tissue. C) Pathways enriched in non-immune regions of the tissue. D) Pathways enriched in immune regions of the tissue.

### Integrative Analysis of single patient derived multi-omics data

One of the primary motivations of multi-modal analysis is the provision of orthogonal data from the directly sampled tumor environment. We therefore performed cross integration of multi-omics data to gain insight into the salient features of the TME. Integrative data analyses were performed on spatial proteomics and transcriptomics, combined single cell RNA sequencing and metabolic profiling of tumor and immune populations, and correlative studies between immunopeptidome profiling and bulk and single cell RNA sequencing results.

#### Core sampling locations

The core sampling locations and analyses are specified in Figures 6A and 6B. The MR coordinates were obtained via neuro-navigated biopsy needles, with 3D coordinates recorded intra-operatively then re-mapped onto patient’s pre-operative scan to confirm correlate sampling locations to their respective assay.

**Fig. 6.**
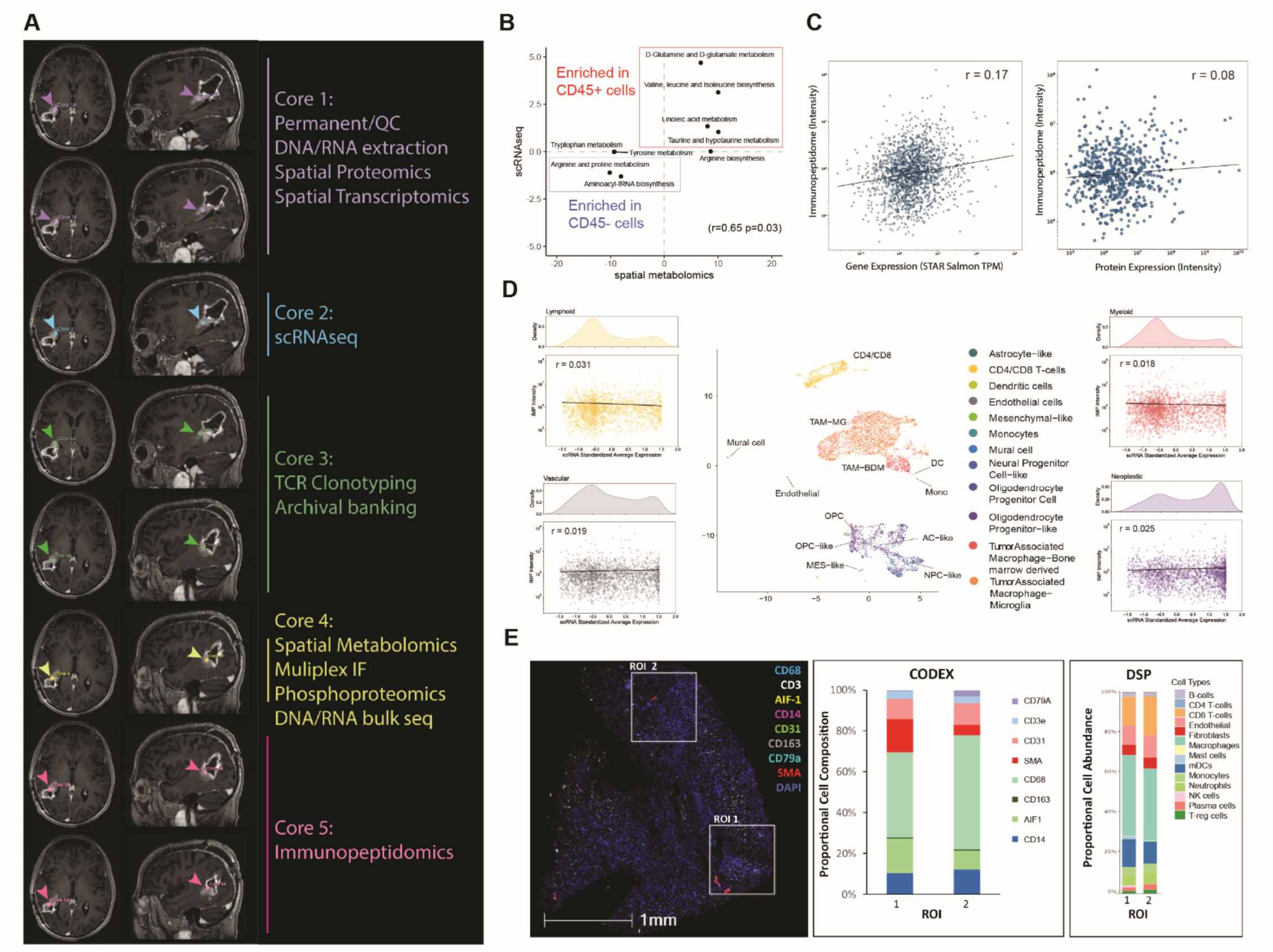
Integrative analysis of single patient derived multi-omics data. A) Core sampling locations and proposed analysis methods. B) Comparison of pathway enrichment analysis of immune vs non-immune cells using spatial metabolomics and scRNAseq. r= Pearson correlation coefficient. C) Correlation between the abundance of MHC peptides and their matching transcripts and proteins. R = Pearson correlation coefficient. D) Correlation of abundance of MHC peptides with corresponding cellular subsets as identified by scRNAseq. (E) Comparison of CODEX protein data to GeoMx-WTA data using corresponding regions of interest. Multiplex immunofluorescence was performed on the biopsied section showing selected ROIs (left panel). Protein expression for phenotypic markers by CODEX (middle) and immune cell deconvolution from GeoMx-WTA (right) showing the distribution of cell type scores of immune cell subtypes in the selected ROIs.

#### Cross-comparison of spatial metabolomics and scRNAseq

Comparison of spatial metabolomics with single cell RNA sequencing data was performed by metabolic pathway enrichment analysis on both datasets. We computed the enrichment in immune and non-immune cell populations and found a moderate correlation (Pearson’s r=0.65, p=0.03) (**Figure 6B**) between the enrichment ratio of selected metabolic pathways. This result indicates agreement between the transcriptomic and metabolomic analyses performed. The top co-nominated pathways for immune cells included D-glutamate metabolism, leucine/isoleucine biosynthesis and linoleic acid metabolism (**Figure 6B**).

#### Correlation between MHC Class I immunopeptidome and bulk and single cell RNA expression

The rank-ordered abundance of MHC peptides assessed by immunopeptidomics did not positively correlate with that of corresponding transcripts (**Figure 6C left**) or proteins (**Figure 6C right**), highlighting the need for immunopeptidomic analysis to directly quantify tumor antigen presentation. Single cell RNA sequencing data generated from tissue from the same procedure allowed us to delve deeper into the different cell types. The abundance of MHC peptides from immunopeptidomics again did not show significant correlation with the corresponding standardized average gene expression across the four major cell types (Myeloid, Lymphoid, Neoplastic, and Vascular cells) detected on scRNA-seq (**Figure 6D**). Although none of the cell types showed strong correlation with the detected immunopeptidome, transcript expression of the ∼1900 source genes did skew towards the neoplastic population, suggesting that the MHC Class I immunopeptidomics data are enriched for tumor associated proteins, however transcript expression is a poor predictor of peptide abundance.

#### Cross-comparison of spatial proteomics profiling with spatial transcriptomics

We used the CODEX and GeoMx -WTA platforms and compared immune cell distribution obtained from similar regions of interest (ROI) (**Figure 6E left panel**) from non-adjacent sections of a single biopsy for quantitative comparison across spatial proteomics and transcriptomics data. Expression of phenotypic markers from CODEX assay (**Figure 6E middle panel)** was comparable with GeoMx-deconvolution data (**Figure 6E right panel)** and indicated comparable immune cell infiltration in the selected ROIs with the predominance of myeloid cells and fewer B and T lymphocytes.

### Estimated intra-core, intra-positional and inter-positional variance between needle core biopsy samples

A critical question to performing multiple needle core biopsies is the underlying sampling variance (*56*). GBM is widely recognized as a highly heterogenous tumor (*57, 58*). We therefore also sought to develop a workflow which would permit cross sample comparisons of needle core histology and copy number changes at the genome level. Specifically, we assessed the variance between 1) intra-trajectory core biopsies – i.e. multiple biopsies taken from the same location within the tumor, and 2) inter-trajectory core biopsies – differences in sample content from distinct trajectories (and hence by extension, regions) of the tumor itself. In order to ascertain the degree of heterogeneity between different core samples, we performed needle core biopsies from spatially distinct locations in a separate patient with recurrent GBM undergoing surgical resection of recurrent tumor (**Table S1**). A total of 7 cores were obtained from 3 needle passes (referred to as regions) of an en-bloc resected recurrent GBM specimen (**Figure 7A**). Each core sample was reviewed by an expert neuropathologist (TB) and tumor regions annotated to obtain an estimated tumor content. Samples from each region were morphologically similar to each other, with the vast majority of regions characterized as tumor interspersed with reactive cellular changes (**Figure 7F**) with region 1 resembling infiltrating margins, and region 2 showing extensive necrosis and calcification. There was, however, a difference in cellularity between sampled locations in region 3. In addition to morphological analysis, the remainder of each core was processed for DNA extraction and low pass whole genome sequencing was performed to identify copy number alterations.

**Fig. 7.**
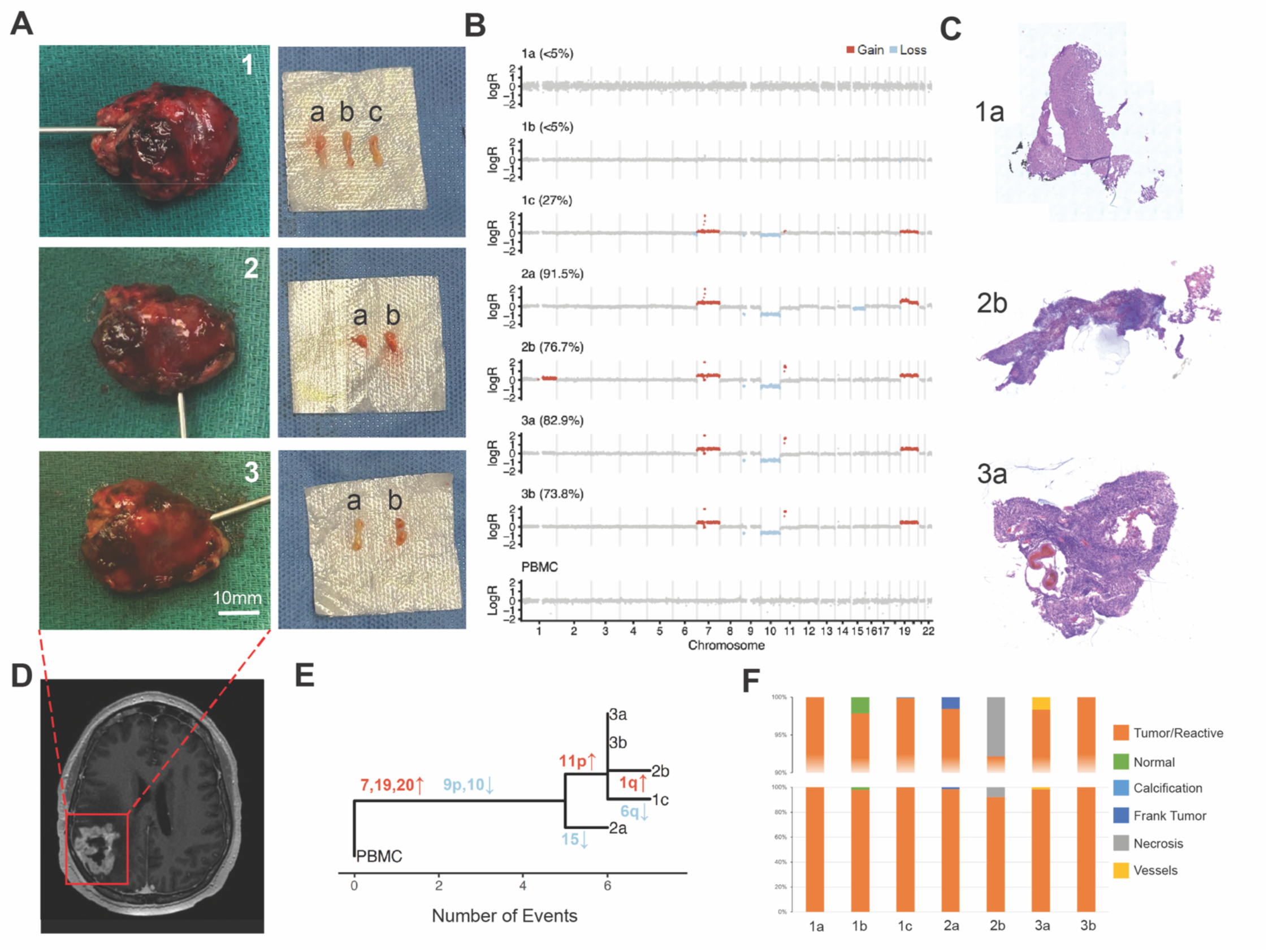
Multi-regional sampling variance analysis. A) A recurrent GBM in resected en-bloc and samples are taken from 3 separate regions of the tumor (labelled 1-3). B) CNV analysis shows low tumor purity in samples 1a and 1b, PBMC control sample is also included in the last track. C) representative H&E sections from each core. D) Pre-operative T1 contrast enhanced and T2 image showing the location of the lesion. E) Phylogenetic analysis demonstrating common shared events such as chromosome 7 gain and 10 loss followed by branching differences separated by sample of origin. F) Histological features by percentage as contoured by a neuropathologist (TB). Note the majority of the specimens’ appearances are non-specific showing mixture of tumor/reactive changes.

Tumor purity was initially evaluated by determining the tumor fraction leading to integer copy number states for gain of chromosome 7 and loss of chromosome 10. Both of these chromosomal alterations are prevalent in GBM and were presumed to be clonal. Tumor purity was initially evaluated by determining the tumor fraction leading to integer copy number states for gain of chromosome 7 and loss of chromosome 10. Both chromosomal alterations are prevalent in GBM and were presumed to be clonal. Samples 1a and 1b were noted to have <5% tumor purity, and were therefore excluded from the downstream analysis, whereas the remainder of the samples had tumor purity ranging from 26-88% purity (**Figure 7B**). Peripheral blood mononuclear cells (PBMCs) from the same patient were included as normal control. Phylogenetic analysis demonstrated that most events were shared across all samples such as gains on chromosomes 7, 19, 20 (including focal amplification of EGFR), as well as loss on 10 and a focal loss on 9p around the CDKN2A locus. Samples had one to two additional subclonal events such as a focal gain on chromosome 11p (shared between 1c, 2b, 3a and 3b) and sample specific events such as loss of 15 in 2a, gain of 1q in 2b and loss on 6q in 1c. Differences within trajectories were pronounced between 2a and 2b, whereas 3a and 3b appear indistinguishable at the copy number level. On the whole, the similarities between samples overshadowed the differences, as samples consistently exhibited a core karyotype comprising alterations on 5 chromosomes, accompanied by a maximum of 1-2 subclonal alterations.

### Generation of patient derived xenografts using needle core biopsy samples

The lack of effective therapeutics in GBM has led to the development of patient derived models to improve our ability to test therapeutics and serve as a platform for the emerging field of precision medicine. Prior studies have shown that small tissue samples from several cancer types are sufficient for generating enough cells to create patient models such as 2D cell lines, 3D organoids/neurospheres or in vivo PDX models (*59–61*). However, the specific success rates for such models in the challenging context of necrotic/reactive tissue available in recurrent glioma stereotactic biopsies is not known.

We retrospectively examined data from model generation we had performed in parallel to a recently reported clinical trial (NCT03152318, to conceptually evaluate the feasibility of planning around simultaneous multimodal data generation and patient model generation from a single neurosurgical procedure **(Figure 8**), (*62*)). Thirty patients underwent stereotactic biopsy to confirm pathologically the presence of high-grade glioma (HGG, WHO grade 3 or 4) on frozen section and in total up to 6 core biopsy samples per patient were obtained. Patients included 28 IDH-wildtype glioblastoma (WHO grade 4), 2 IDH-mutant astrocytoma (1 WHO grade 3 and 1 grade 4) and had the expected spectrum of genomic and clinical characteristics for these diagnoses. Neuropathologist-estimated composition of each biopsy revealed a variable degree of tumor and necrosis with tumor content ranging from 5-80% and all cores having 30% necrosis or less. Transcriptional silencing of the O6-methylguanine-DNA-methyltransferase (MGMT) gene was inconsistent among the group, with 9 patients (30%) demonstrating partial or full MGMT promoter methylation. Next-generation targeted exome sequencing of these gliomas demonstrated a broad landscape of canonical genomic alterations, with particularly high frequencies observed for EGFR and CDK6 copy number gain as well as copy number loss of PTEN and CDKN2A **(Figure 8C)**.

**Fig. 8.**
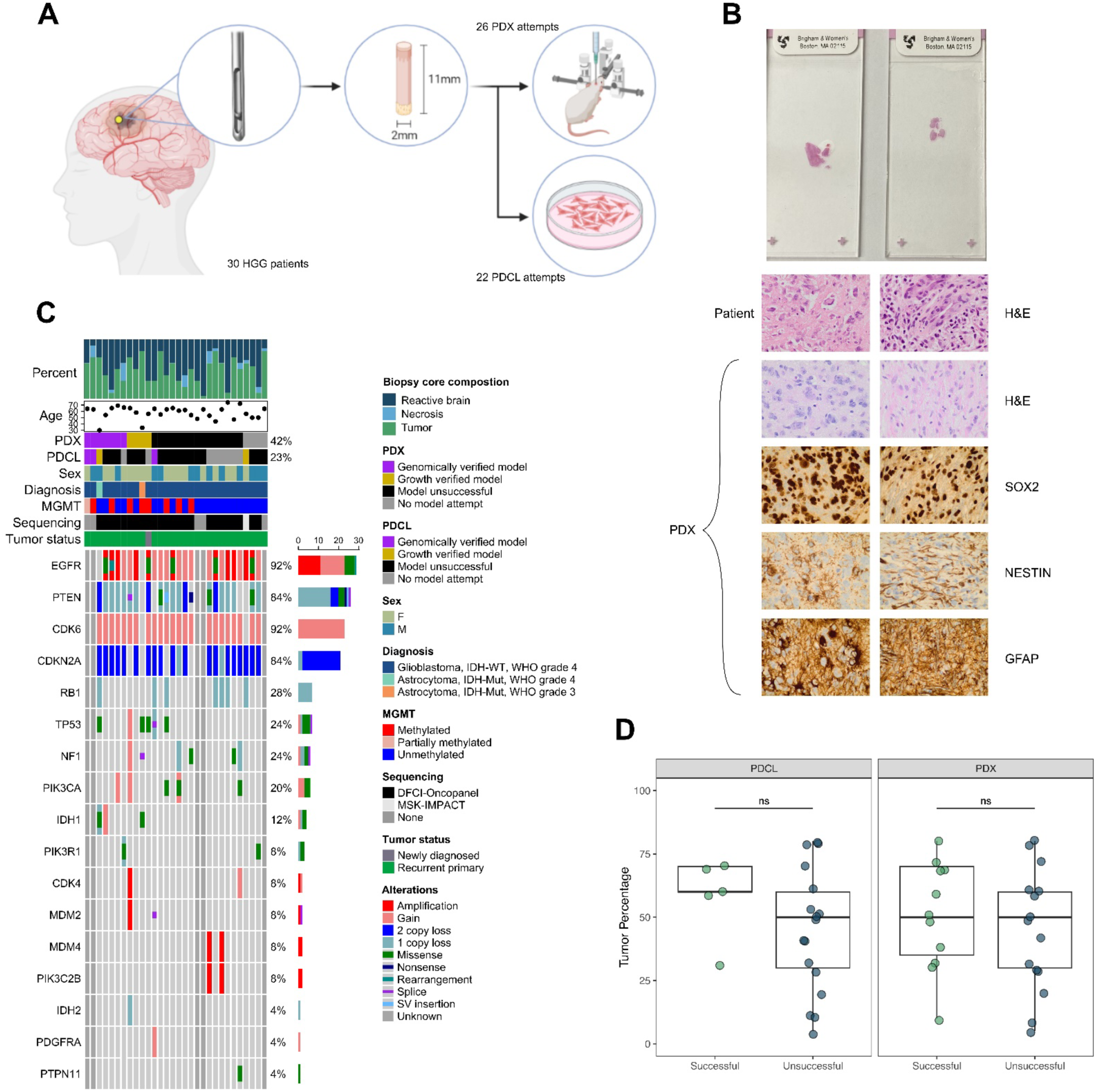
In vivo and in vitro modeling of glioblastoma from stereotactic needle core biopsy. A) Stereotactic needle core biopsies were obtained from 30 HGG patients and used in 26 PDX and 22 PDCL attempts. B) Representative tissue sections from two patients demonstrate the small size of stereotactic needle biopsy cores and microscopic glioma histology, as well as clear evidence of PDX glioma histology via positive immunohistochemical staining for clinically reliable glioma markers. C) 42% (11/26) of PDX attempts and 23% (5/22) of PDCL attempts were successful for a cohort of patients that demonstrated a diversity of histopathological, clinical, and tumor genomic characteristics. D) Neuropathologist-estimated tumor percentage in biopsy samples did not significantly influence model generation success.

A single core biopsy was used to attempt model generation. Cores were dissociated to single cells and sufficient cells were used to attempt different model types depending on number of cells retrieved. Cells were either directly implanted into the brain orthotopically (PDX), or concurrently cultured in vitro as 3D neurosphere patient derived cell lines (PDCL) as previously described (*63*). We attempted both orthotopic PDX and PDCL (n=18 patients), PDX only (n=8) and PDCL only (n=4) in 30 total HGG patients. In total we performed 26 PDX attempts and 22 PDCL attempts. Growth-verified successful PDXs (defined as histologically verified tumors within the brain) were generated from 11/26 patients (42.3%) which is analogous to the maximal success rates reported (*64*). Successful PDCLs were generated from only 5/22 patients (22.7%) which was lower than that generally seen for in vitro neurosphere generation (*65*). Growth verified models were obtained even from samples with lower tumor content or significant necrosis with the specific tumor content or necrosis within the range. All successful models demonstrated clear features of diffuse high grade glioma histology by hematoxylin & eosin (H&E) staining as well as lineage verification by immunohistochemistry for glioma defining markers including SOX2, Nestin, and GFAP **(Figure 8B)**.

We also sought to determine if a sample’s estimated tumor percentage influenced the likelihood of model generation in a meaningful way. Successful PDX models were generated from tumors ranging between an estimated 10-70% tumor content and 0-30% necrosis. Successful PDCL models were generated from tumors ranging between 30-70% tumor content and 0-20% necrosis. Statistical analysis by Student’s t test demonstrated that tumor percentage did not yield a significant influence over model success in our cohort for both PDX (p=0.5168) and PDCL (p=0.1919) attempts, though there appears to be a trend toward significance in the PDCL cohort **(Figure 8D)**.

In summary, both in vitro and in vivo models were readily generated from stereotactic core biopsies but with higher success rates for in vivo PDX. Tumor content did not significantly influence model generation success, though the small size of our cohort warrants further investigation into this topic.

## Discussion

GBM remains the most common and lethal primary brain cancer with minimal incremental improvements in patient outcomes despite over a century of research and trials. It is now understood that GBM has a highly immunosuppressive, immune “cold” tumor microenvironment, impervious to current therapeutic efforts. The lack of progress in GBM therapeutics can be partially attributed to the lack of understanding of disease response and adaptation under therapeutic pressure. This lack of visibility into how cellular states alter and respond to cytotoxic stress, how tumor-immune and tumor-stromal interactions interact to generate an immunosuppressive tumor milieu that favors progression and resists treatment, has handicapped efforts to design more rational therapies.

Diagnostic needle core biopsies have been the mainstay of neurosurgical diagnosis for decades and the indications, safety profile and histological diagnosis that can be obtained from core biopsies are well established (*8*). However, the use of biopsies in the context of gaining additional information pertinent to a patient’s disease course beyond diagnosis, what we term investigative biopsies, has not been established. The central premise of this study is that standard needle biopsy cores contain a wealth of information which at present is underutilized. Possible reasons for this include concerns about undue risk to patients, technical challenges in obtaining high resolution data, and the lack of experience in generating multi-omics data from low volume samples. These reasons result in a lack of rationale and interest in investigative sampling, despite a clear need.

The ability to generate high resolution, data rich tissue profiling data to gain an understanding of a patient’s disease course potentially alters the risk-benefit calculus for investigative biopsies. Extending this concept further, longitudinal investigative biopsies may permit unprecedented insight into tumor and immune responsiveness to therapy. Underpinning this entire concept, however, is the need to establish the feasibility of obtaining high quality multi-modal measurements from a single patient in the same procedure.

Large scale proteogenomic approaches integrating genomic data with proteomics and post-translational modifications such as protein phosphorylation such as those by the National Cancer Institute Clinical Proteomic Tumor Analysis Consortium (CPTAC) have revealed new biological mechanisms and ushered in new integrative computational methods (*66–71*). The advent of single cell profiling has the potential to refine these associations further with corresponding spatial correlations at the intraoperative macroscopic and microscopic tissue level scale. Multi-omics analysis of GBM tumors has previously identified core transcription factors associated with mesenchymal transformation in GBM (*72*). Proteogenomic analyses have found cellular stress and growth factor networks regulating the EGFR signaling pathway (*73, 74*). A similar stress response phenomenon is also seen when integrating metabolic and proteomic profiles (*75*). Recently, Dekker et al. performed gene expression, proteomics and phosphoproteomics analysis on 8 primary and recurrent sample pairs and reported that differences found at the transcriptional level were not seen at the proteomic level, despite more pronounced differences between primary and recurrent samples seen in the proteomics data (*76*). This data reinforces the view that proteomics data provides orthogonal information which needs to be integrated into the overall picture of the TME. It should be noted however, that in all studies, the specimen type was not specifically sourced from needle core biopsies, which remain the most abundant and accessible specimen type in routine clinical practice. The ability to perform integrative multi-omics analysis on needle core biopsies alone therefore has significant implications on the availability of tissue specimens for study. Single cell analysis of needle core biopsies in brain tumors has been attempted in the past however the number of reported cells sequences have been typically low (*30, 31*), with the number of cells per biopsy ranging between 70-400 cells per core. With advances in tissue processing and single cell encapsulation, we have shown that pre-sequencing cell numbers in the region of 250,000-600,000 cells can be generated from 2 cores, which represents a difference of several orders of magnitude, and well within the range of standard requirements for single cell RNA sequencing. Remarkably, these cellular yields were obtained from previously treated, recurrent tissue, where one would expect lower cell yields, affirming the robustness of the protocol, standing in contrast to previous reports which were obtained from newly diagnosed cases (*30, 31*). Besides scRNA, we also demonstrate an unprecedented breadth of information which can be obtained from standard needle biopsy cores through various assays. Although multi-core sampling is part of routine surgical practice, the application of systematic sampling for targeted multi-omics analysis combined with precise geotagging of sampling coordinates is uncommon and provides additional macroscopic spatial information (**Figure 6A**). The analysis techniques were chosen based on the organizing principle of tumor-immune co-evolution. Modalities were specifically selected to resolve cell type composition and cell states, spatial cellular profiles at both the gene expression and proteomic level, intra-tumoral TCR clonal frequencies sequences (**Supplemental Figure 1**), buttressed by orthogonal phosphoproteomics data, spatial metabolomics data and intra-tumoral antigen diversity via MHC Class I immunopeptidomics, all obtained via minor modifications to a standard neurosurgical procedure. The data generated is of high quality and relevance, and the results of this study stand as a proof of principle that needle core biopsies can serve as suitable input material for high resolution downstream analyses.

We also performed integrative analyses across modalities in addition to demonstrating feasibility of multi-omics technologies. These cross-cutting analyses provided insight into the metabolic states of specific immune and tumor cell types. It was, for example, noted that the linoleic acid pathway was upregulated in CD45+ regions of the TME. We were also able to cross correlate spatial proteomics imaging and whole transcriptome regions of interest, highlighting the utility of these interrelated modalities. Immunopeptidomics assays also allowed us to correlate the presented HLA Class I peptidome at the bulk and single cell RNA expression level, and in concordance with previous studies, show that gene expression correlated poorly with the detected surface peptides (*77, 78*), again highlighting the need for orthogonal multi-omics measurement. Having this wealth of data will also assist in efforts to develop peripheral biomarkers. Future efforts will include concurrent analysis of sample blood and CSF, with the goal of identifying peripheral biomarkers of therapeutic or immune response to treatment which could obviate the need for repeated tissue sampling. Finally, from a translational standpoint stemming from a need to create patient specific tumor models, needle core biopsies often contain sufficient tissue to generate patient derived xenograft (PDX) models, with 42% of needle cores going on to generate viable tumors in immuno-compromised mice. Somewhat counter-intuitively the success rate in generating in vivo tumor avatars was higher than the success rate in generating cell lines from tissue, which may be related to the conduciveness of the initial seeding environment.

In conclusion, we have demonstrated that multiple needle biopsies taken from a single procedure can be used to generate high resolution cellular and molecular information regarding immune cell content, gene expression profiles, and activated protein pathways and antigenic diversity. The natural progression for this work would be to extend this to additional timepoints. It is anticipated the value and utility would be greatly enhanced through longitudinal, serial sampling from the same patient. Efforts are currently underway aimed at expanding the use of investigative biopsies in a clinical trial setting.

## Materials and Methods

### Experimental Design

This is a single subject experimental design where multi-modality measurements are taken from the tissue samples from the same subject. For the PDX generation section, the experimental design is that of a retrospective correlative study examining tissue from a cohort of patients who were enrolled in a prior clinical trial arm.

### Ethics Approval

Tissue collection and material transport were governed under local institutional IRB (MSKCC #06-107 & #09-156, DF/HCC #10-417 & #16-557). Written patient consent was obtained prior to surgery in all cases.

### Sample collection, geotagging and transportation

Biopsy cores were collected immediately during operation and transported in MACS storage solution (Miltenyi, Germany). Samples were then sent to the primary processing institution (MDACC) on ice via courier.

### Sample Distribution

Formalin fixed samples were processed locally at MSKCC (Core 1). Fresh tissue placed in tissue storage solution and was immediately shipped for central processing at MDA (Core 2 & 3). Snap frozen specimens (Cores 4-5) were shipped to BWH/DFCI and MIT respectively.

### Low volume glioma tissue processing

At the receiving institution, tissue cores were processed within 24 hours of the operation. Tissue cores were morselized into small pieces under sterile conditions using sterile scalpels, triturated using pipettes then transferred to a 50ml falcon tube (Corning, USA), then incubated with papain (37°C, 25 min)(Worthington, USA) according to manufacturer instructions. After incubation HBSS without magnesium or calcium was then added to make up to 50ml and the sample centrifuged (300×g, 5 min, 4◦C), supernatant aspirated before being passed through a 70μm cell strainer (BD, USA). Erythrocytes were removed by resuspending and incubating the obtained pellet in 5 ml of ACK-lysis buffer (eBioscience, USA) for 10 min, followed by centrifugation (300×g, 5 min,4°C ). Residual myelin and extracellular debris was eliminated using the Debris Removal Kit (Miltenyi Biotech, Germany). Cell counts were quantified using an automated cell counter (Nexcelom, USA) after resuspending the pellet in PBS. Cells were stored in Bambanker (Wako, Japan). Cell suspensions were immediately placed in a storage container (Mr. FrostyTM, Thermo Fisher Scientific, USA) and stored in LN2.

### Liquid chromatography tandem mass spectrometry (LC-MS/MS)

For pTyr proteome, the enrichment of pTyr peptides was performed by incubating digested peptides with 60 μl of protein G agarose beads (Sigma) pre-conjugated to 24 μg of 4G10 (Bioxcell) and 6 ul of PT66 antibodies overnight at 4 °C in immunoprecipitation (IP) buffer (100 mM Tris-HCl, 1% NP-40, pH 7.4). Subsequently, immunoprecipitated peptides were eluted twice with 0.2% trifluoroacetic acid (TFA) for 10 min each and enriched for phosphopeptides using ferric nitrilotriacetate (Fe-NTA) columns (Thermo). Eluates from the Fe-NTA columns were dried using vacuum centrifuge, reconstituted in 4ul of 3% acetonitrile/0.1% formic acid, and directly bomb-loaded onto a hand-packed 10 cm analytical column containing 3 μm C18 beads. Agilent 1100 Series HPLC connected to Orbitrap Exploris 480 Mass Spectrometer was operated at 0.2 ml/min flow rates with a precolumn split to attain nanoliter flow rates through the analytical column and nano-electrospray ionization tip. Peptides were eluted with the increasing concentrations of buffer B using the following 140 minute gradient settings (Buffer A: 0.1% acetic acid; Buffer B: 70% acetonitrile/0.1% acetic acid): 0 min: 0% B; 10 min: 11% B; 110 min: 32% B; 125 min: 60% B; 130 min: 100% B; 128 min: 100% B; 130 min: 0% B. The MS parameters were: ESI spray voltage, 2.5 kV; no sheath or auxiliary gas flow; heated capillary temperature, 275 °C, data-dependent acquisition mode with the scan range of 380–2000 m/z. MS1 scans were acquired at 60,000 resolution, maximum injection time of 100 ms, normalized AGC target of 300%, and only included the precursor charge states of ≥ 2 and ≤ 6. For every full scan, MS/MS spectra were collected during a 3 seconds cycle time. For MS/MS, Ions were isolated (0.7 m/z isolation width) for the maximum IT of 250 ms, normalized AGC target of 100%, HCD collision energy of 30% at a resolution of 60,000, and the dynamic exclusion time of 35 seconds. Half of the supernatants from pTyr-IP were subjected to high-pH reverse-phase fractionation on a Kromasil® C18 HPLC Column (5 μm particle size, pore size 100 Å, L × I.D. 250 mm × 4.6 mm) using buffer A (10 mM triethylammonium bicarbonate (TEAB), pH 8) and buffer B (10 mM TEAB, pH 8, 99% acetonitrile) over an 85-minute gradient (0 min: 1% B, 1 min: 1% B, 5 min: 5% B, 65 min: 40% B, 75 min: 70% B, 84 min: 70% B, 85 min: 1%) into 10 fractions (concatenated) using a Gilson FC 204 Fraction Collector. Fractionation was performed at a flow rate of 1ml/min and 1 min per fraction between 10-85min portion of the gradient. For each fraction, 1/10 was allocated for protein expression analysis and 9/10 was allocated for global phosphoproteomic (pSer/Thr) analysis. Fractions for pSer/Thr analysis were resuspended in 50ul of 0.2% TFA and subjected to Fe-NTA column-based phosphopeptides enrichment, after which they were eluted, dried, and resuspended in 3% acetonitrile/0.1% formic acid for injection by Dionex UltiMate 3000 Autosampler (Thermo).

For immunopeptidome, MHC-peptide complexes were isolated by IP (anti-HLA-A/B/C; clone W6/32 (BioXcell); conjugated to 20 μl FastFlow protein A sepharose bead (GE healthcare)), eluted with 10% acetic acid, filtered by size exclusion filter (Nanosep 10K, PALL), cleaned up with zip tip (Thermo), and dried with speed-vac. Samples were resuspended in 3% acetotnitrile/0.1% formic acid and directly loaded onto an analytical capillary chromatography column (about 1 μm orifice) prepared and packed in-house (50 μm ID × 15 cm and 1.9 μm C18 beads, ReproSil-Pur). The sample was analysed using an Orbitrap Exploris 480 Mass Spectrometer (Thermo Fisher Scientific), coupled with an UltiMate 3000 RSLC Nano LC system (Dionex), Nanospray Flex ion source (Thermo Fisher Scientific) and column oven heater (Sonation; operated at 45°C). Peptides were eluted using a 125 minute gradient (Buffer A: 0.1% formic acid, Buffer B: 80% acetonitrile/0.1% formic acid) with the following minute:B% profile: 0:3, 30:3, 31:8, 85:30, 97:50, 100:97, 103:97, 103.1:3. The flow rate of 0.4 μl/min was used between 0-29.5 min and 0.075 μl/min between 30-125 min. The MS parameters were: spray voltage, 2.5 kV; no sheath or auxiliary gas flow; heated capillary temperature, 275 °C, data-dependent acquisition mode with the scan range of 350–1,200 m/z. MS1 scans were acquired at 60,000 resolution, maximum injection time of 50 ms, standard AGC target, and only included the precursor charge states of ≥ 2 and ≤ 4. For every full scan, MS/MS spectra were collected during a 3 seconds cycle time. For MS/MS, Ions were isolated (0.7 m/z isolation width) for the maximum IT of 250 ms, normalized AGC target of 100%, HCD collision energy of 30% at a resolution of 60,000, and the dynamic exclusion time of 30 seconds.

### Mass spectrometry data analysis

Mass spectra were analyzed using Proteome Discoverer (v3.0, Thermo Fisher Scientific) and searched using Mascot (v2.8) against the human Swiss-Prot database (v2022_11). For peptide bound HLA, peptides were searched with no enzyme and variable methionine oxidation. Peptide spectrum matches were filtered by an ion score ≥15, length 8-11, search engine rank of 1, and aggregated across unique peptides. GibbsCluster 2.0 was used for motif analysis (*79*) and the figures were generated by Seq2Logo. For phosphoproteomic data, raw files were searched with two or fewer missed cleavages, precursor and fragment ion matched with a 10 ppm and 20mmu mass tolerances, and fixed modifications of carbamidomethyl (Cys) and dynamic modifications of phosphorylation (Ser, Thr, Tyr) and oxidation (Met). After quantification by minora feature detection method, peptide spectrum matches were filtered by an ion score ≥20 and search engine rank of 1. The ptmRS node (*80*) in Proteome Discoverer was used to confidently assign the phosphorylated and oxidized sites (filtered for peptides with localization probability ≥ 90% after search). All data were processed in R studio (v4.1.0).

### Single-Cell RNA Sequencing

scRNA-seq was performed using the 10x Genomics Chromium Single Cell Controller. Single-cell suspensions were prepared from tissue biopsy as described above. Freshly isolated cells were droplet-separated using the Chromium Single Cell 3′ v.3 Reagent Kit (10x Genomics, USA) with the 10x Genomics microfluidic system creating cDNA library with individual barcodes for individual cells. Barcoded cDNA transcripts from patients were pooled and sequenced using the NovaSeq 6000 Sequencing System (Illumina, USA).

### Single-Cell RNA sequencing analysis

Raw single-cell RNA seq reads generated by Illumina sequencer were demultiplexed into FASTQ and aligned to GRCh38 reference genome to generate count matrices using Cell Ranger v6.0.0 analysis pipelines (10x Genomics). The preliminarily processed data was used for downstream analysis by Seurat v4.3.0 (*81*). Low-quality and dead cells which had <200 or >2,500 unique transcripts or >5% of mitochondrial transcripts were filtered out. Multiple datasets (with and without inhibitors) were combined, normalized, identified variable features, and applied principal component analysis (PCA) using Seurat. Batch effect correction of combined data was performed using Harmony algorithm v0.1.1 (*82*). Following batch correction, FindClusters was applied with a resolution of 0.5 to identify cell clusters and uniform manifold approximation and projection (UMAP).

### Spatial proteogenomics

Digital Spatial profiling (DSP, Nanostring) experiments were performed according to the manufacturer’s protocol and as previously described (Beechem JM, 2020, PMID: 31502169). Briefly, slides were stained and imaged to visualize morphology markers, GFAP-AF488, 53-9892-82, Clone GA5, diluted 1-100, Syto83, S11364, diluted 1:25, CD45-AF594, NBP1-44763AF594, clone EM05, diluted 1:100, Ki67-AF647, NBP2-22112AF647, clone 8D5, diluted 1:100. Images at ×20 magnification were assembled to yield high-resolution regions of interest. As previously described, ROIs were illuminated and released tags were collected into 96-well plates. Sequential sections were used for GeoMx Protein and RNA profiling. For protein detection GeoMx (v1.0) Human NGS Protein Core, (v1.0) Human NGS Cell Death Protein, (v1.0) Human NGS Glial Cell Subtyping Protein, (v1.0) Human NGS Immune Activation Status Protein, (v1.0) Human NGS Immune Cell Typing Protein, (v1.0) Human NGS IO Drug Target Protein, (v1.1) Human NGS MAPK Signaling Protein, (v1.0) Human NGS Myeloid, (v1.0) Human NGS Neural Cell Typing Protein, (v1.0) Human NGS Pan-Tumor Protein, (v1.0) Human NGS PI3K/AKT Signaling modules were used to identify cell types. Detection of mRNA was performed for morphology marker– defined compartments according to the NanoString GeoMX RNA assay protocol described previously (*83*) using the Whole Transcriptome Atlas probe reagent. Library preparation was also performed according to the manufacturer’s protocol. DSP collection sample plates were dried, resuspended in nuclease-free water, and amplified using PCR according to the manufacturer’s protocol. Purified libraries were sequenced by Illumina NovaSeq6000. The FASTQ reads from sequenced DSP library were processed by the GeoMx NGS DnD Pipeline to convert sequencing reads into counts per ROI (NanoString, MAN-10133_03). After processing, counts were uploaded to the GeoMx DSP Data Analysis Suite (NanoString Technologies, USA).

### CODEX

A 4 µm FFPE tissue section was placed on a positively charged glass slide and stored at 4°C until use. Codex staining assay was performed following the manufacturer’s protocol. Purified antibodies, barcodes and reporters were purchased commercially as listed in Supplementary Table 4. Antibodies CD1c, Bcl-6, DC-LAMP, CXCL13, CXCR5, and CD206 were conjugated through the Spatial Tissue Exploration Program (STEP) from Akoya Biosciences. For GFAP, Olig2, FoxP3, CCR7, Lag3, CD138, and AIF1, purified antibodies (free of BSA and glycerol) were conjugated in-house. The remaining antibodies were commercially tagged with PhenoCycler® Barcodes at the time of purchase from Akoya Biosciences. The in-house antibody conjugation process followed the manufacturer’s guidelines. Purified barcoded-conjugated antibodies were used within 6 months of conjugation.

#### Codex imaging, processing, segmentation, and analysis

The stained tissue section was captured using PhenoCycler-Fusion (version 1.0.8) using DAPI, ATTO550, CY5 and AF750 filters at a scan resolution of 0.50um (20X) and saturation protection was applied on DAPI channel. Alignment of images across cycles, stitching of tiles and subtraction of auto-fluorescence was performed using CODEX® Processor application. Cell segmentation was performed by applying StarDist, a deep learning-based algorithm, to the DAPI channel to segment nuclei; cells were approximated based on expansion from the nuclei. Scripts for performing the segmentation in the QuPath software and a pre-trained model were provided by Akoya Biosciences. All mIF analyses were run in R-4.2.1.

#### Cell clustering

The cell expression values were first Z-score normalized across markers followed by application of scaled and centered biomarker expression value for each cell.(Seurat citations). Principal component analysis was performed to identify 20 principal components (“Seurat::RunPCA, npcs = 20”), which were then used in the Harmony algorithm to group cells and correct for dataset-specific and experimental conditions (“harmony::RunHarmony”) (*81, 84*). The Harmony reduction was used for downstream analysis. The dimensionality was reduced with UMAP (“Seurat::RunUMAP, reduction = ‘harmony’”). Clustering was performed using a shared nearest neighbor modularity optimization-based algorithm. The UMAP plot and a heatmap plot showing the average normalized marker expression in each cluster were graphed using the Seurat package. Clusters were annotated using their average expression to identify cell types, and these annotations were validated by manual inspection of multiplexed IF stains on images. The cluster spatial location plot was graphed using the ggplot2 package.

#### Cell neighborhood analysis

To define cellular neighborhoods (CNs), the nearest 20 neighbors for each cell were recorded and set as a window. The number of each cell cluster in the window for each cell was counted, resulting in a matrix of cells by cell clusters, with each row representing a cell, each column representing a cell annotation (cell type) from the clustering above, and each value representing the count of neighbors of the given annotation. The neighbor cell proportion was computed for each row. The resulting matrix was clustered using k-means clustering (“stats::kmeans”), where the optimal k was determined empirically by maximizing the silhouette score metric (“factoextra:: fviz_nbclust”). Each cluster was defined as a CN. Thus, each cell was given both a cell type annotation, which depends only on the cell’s own marker expression, and a cell neighborhood annotation, which depends on the cell’s type and the identities of its nearest neighbors.

### MALDI-MSI tissue preparation and imaging

Biopsy Core Four underwent a preservation process, being rapidly frozen on dry ice. For MALDI MSI the core was then cryo-sectioned into 10 µm-thick slices. These slices were subsequently thaw-mounted onto an indium tin oxide (ITO) slide. Serial sections were prepared for microscopy staining. For the mass spectrometry analysis, we utilized a 1,5-diaminonaphthalene hydrochloride MALDI matrix, with 15N glutamate spiked in as an internal standard. The matrix was prepared at a concentration of 4.3 mg/mL in a mixture of 4.5 parts HPLC grade water, 5 parts ethanol, and 0.5 parts 1 M HCl (v/v/v). A TM-sprayer from HTX Imaging was used to spray the matrix, with the following parameters: a flow rate of 0.09 mL/min, spray nozzle velocity of 1200 mm/min, spray nozzle temperature of 75 °C, nitrogen gas pressure of 10 psi, track spacing of 2 mm, and a four-pass spray. The mass spectrometry analysis was carried out using a 15 Tesla SolariX XR FT-ICR MS (Bruker Daltonics, Billerica, MA). The instrument was set to negative ion mode, and the mass range scanned was from m/z 46.07 to 3000, with a sampling step size of 30 µm. Each sampling point consisted of 200 laser shots at a laser power of 21% (arbitrary scale), with a laser repetition rate of 1,000 Hz. We employed the Continuous Accumulation of Selected Ions (CASI) mode, setting Q1 to m/z 150 with an isolation window of 200. The mass range was calibrated using a tune mix solution from Agilent Technologies with the electrospray source. Additionally, the internal standard 15N glutamate was used for on-line calibration during the acquisition. For data analysis, SCiLS Lab software (version 2023c Pro, Bruker Daltonics, Billerica, MA) was used to view and process ion images and mass spectra. The dataset was normalized to the total ion current (TIC), and peaks were annotated using Metaboscape (2021b,Bruker Daltonics, Billerica, MA)). Metabolites were putatively annotated based on an accurate mass with a Δppm < 0.5 and MSMS measurements. MetaboanalystR (*85*) was used to perform pathway enrichment analysis at every pixel. The resulting enrichment ratios were displayed spatially with an in-house R script. Metabolite differential expression was tested with a t-test and FDR correction.

### Tissue-Cyclic Immunofluorescence (t-CyCIF)

Tissue-based cyclic immunofluorescence (t-CyCIF) (1) of fresh frozen tissue was adapted from a detailed protocol available at protocols.io (dx.doi.org/10.17504/protocols.io.bjiukkew). Fresh frozen tissue samples mounted on SuperFrost slides (Fisher Scientific #12-550-15, Hampton NH) were fixed with 4% PFA for 1 hour at room temperature followed by membrane permeabilization for 5 minutes in 0.5% Triton X in PBS (Thermo Scientific #28316). All primary and conjugated antibodies were incubated overnight at 4°C in the dark. Secondary antibodies targeted to unconjugated primary antibodies in the first cycle were incubated for 1 hour at room temperature. Hoechst 33342 co-staining was performed by combining Hoechst with primary antibodies in SuperBlock Blocking buffer (Thermo Scientific #37515) buffer at every cycle. Coverslips were mounted on slides with 10% Glycerol in PBS prior to imaging. Following imaging, coverslips were removed in 1X PBS, then fluorophores were inactivated by incubating slides in a solution of 4.5% H_2_O_2_, 24mM NaOH in PBS for 1 hour under an LED light source. Slides were processed through multiple cycles of antibody incubation, imaging, and fluorophore inactivation. CyCIF images were acquired on a CyteFinder II slide scanning fluorescence microscope (RareCyte Inc. Seattle WA) with CyteFinder software (v3.11.024). t-CyCIF image processing and data quantification was performed as previously described (2). Stitching and registration of tiles and cycles were done in MCMICRO (3) using ASHLAR (v1.10.2) (4) module. Code is available on Github (https://github.com/labsyspharm/cd73_coy_spatialcorrelation) and Zenodo (https://zenodo.org/record/6628875#.YqJVfqjMJD8). DOI:10.5281/zenodo.6628875. Additional details and code can be found at www.cycif.org. A list of antibodies used in CyCIF is available in Supplementary Table 5.

### MALDI-MSI and t-CyCIF integration

t-CyCIF images were registered onto the MSI space using an in-house MATLAB scipt. Several fiducials were specified manually around the contour and specific tissue landmarks. An affine transformation was obtained from the points using the MATLAB function fitgeotrans. The obtained transformation was applied to all CyCIF channels.

### MALDI-MSI and scRNAseq integration

Metabolite pathway enrichment analysis was performed using MetaboAnalyst (*86*) for spatial metabolomics data and PANTHER (*87*) for scRNAseq data. The enrichment based on spatial metabolomics data was used to select a list of top 5 pathways enriched in each group (immune cells vs non-immune cells). Due to mismatching names in the corresponding libraries (KEGG (*88*) and GO (*89*) the crosslink was performed manually.

### PDCL and PDX Generation

Patient samples for model generation DF/HCC protocol #10-417 from patients who planned to enroll to DFCI clinical trial #16-557 NCT03152318 of oncolytic HSV agent CAN-3110. Biopsy cores deemed to be in excess of clinically required tissue were allocated for research use. Fresh tumor biopsy tissue specimens were transported in DMEM + 1% penicillin-streptomycin. Fresh tumor biopsy tissue was dissociated in NeuroCult NS-A media to a single-cell level and viable cells counted. PDX attempts were conducted via intracranial mouse injection per DFCI institutional animal protocol and protections. Mice were sedated using a 0.2 L/minute isoflurane administration along with oxygen and arranged within a stereotactic apparatus. Tumor cells were injected into the right striatum as 100,000 cells in 1uL of NSA media. Mice were subsequently monitored at a minimum of 3 times per week for symptomatology as the endpoint. Histological confirmation of bulk tumor was performed by a neuropathologist (KLL). PDCL attempts were performed by placing a minimum of 100,000 viable cells on laminin using the NeuroCult NS-A Proliferation Kit (StemCell Technologies) supplemented with 0.0002% heparin (StemCell Technologies), EGF (20 ng/mL), and FGF (10 ng/mL; Miltenyi Biotec) in a humidified environment of 5% CO^2^ at 37°C. Models were growth verified once five serial passagings were completed indicating long term growth. Growth verified models were genomically verified via whole exome sequencing for glioma diagnostic copy and mutation aberrations. All models and relevant data are available from the DFCI Center for Patient Derived Models. Tumor genomic alteration data was obtained by targeted next-generation sequencing via DFCI Oncopanel or MSK-IMPACT. These validated measures identify genomic alterations including single-nucleotide variants, insertions and deletions, copy number alterations, and structural variants in the exons and selected introns of 447 (Oncopanel) and 410 (MSK-IMPACT) genes, including genes relevant to the diagnosis and subclassification of HGG. Sequencing results were reviewed and reported by a board-certified neuropathologist. Sequencing data from these assays, as well as clinical information regarding age, trial cohort, diagnosis at time of injection, and MGMT promoter methylation status, was used to generate an Oncoprint genomic profile using R 4.2.2, RStudio 2022.12.0+353, and the Oncoprint function of the ComplexHeatmap 2.14.0 package. Statistical analysis (Student’s t test) evaluating the influence of tumor percentage on model success was conducted using the same version of R (4.2.2) and RStudio (2022.12.0+353).

### Bulk DNA and RNA Sequencing and Analysis

Each core was processed for DNA, RNA, and protein extraction using the Nucleospin Tri-kit (Machery-Nagel). Samples were homogenized first in a 1ml pipette and then 10 times through a 26G needle before being prepared per manufacturer’s recommendations. Extracted DNA and RNA were evaluated by Fragment Analyzer (Agilent) and 1ng of DNA was used for nexteraXT library preparation (Illumina) before low pass CNA whole genome sequencing on either an Illumina NextSeq500 (40nt paired end) or Singular G4 (50nt paired end). Bulk RNAseq libraries were prepared from 10ng of RNA using the Pico-input strand-specific total RNA-seq for mammalian samples v2 kit from Takara. DNA samples were sequenced on an Illumina NextSeq500 (40 paired end). RNA samples were sequenced on a MiSeq to provide validation of the methodology using a 60nt forward read and were analyzed using the RNAseq NextFLow pipeline. DNA reads were mapped to the hg19 reference genome using BWA-MEM. We calculated the number of reads in 500kb genomic bins throughout the genome. In the case of tumor samples, read counts were normalized based on the read counts from the PBMC sample, followed by GC correction using HMMcopy (*90*) to get read depth ratio’s R. These values were then logged to get LogR values plotted in the figure. Gains and losses were identified using QDNAseq (*91*) with a probability cutoff of 0.5. To estimate tumor fraction we calculated the purity that resulted in average purity corrected copy number values of 3 for chromosome 7 and 1 for chromosome 10. Gains on chromosome 7 and losses on chromosome 10 are ubiquitous features in GBM and were assumed clonal. Phylogenetic tree were constructed by generating a binary event matrix where rows were genome segments and columns were samples. Genome segments were defined as continuous regions of the genome that had gains or losses in any sample, we ensured that segment boundaries were identical across samples. Matrix values were 1 if a sample had a copy number alteration in that segment and 0 otherwise. Hamming distances were calculated between all samples and a tree constructed using the neighbor joining algorithm using the distance matrix as input.

## Data Availability

All data produced in the present study are available upon reasonable request to the authors

## Acknowledgements

Spatial proteomics and transcriptomics analysis was performed with the kind assistance of Nanostring Technologies, USA. This study was assisted in part by the MD Anderson CATALYST program. Figures 1,2 and 8 created with BioRender.com

## Author Contributions

Conceptualization: The GBM TeamLab

Methodology: KKHY, SB, GB, MSR, RA, JG, MCP, SJ, AD, MJW, YE, ZA-M, KH, SWM, AS, FFL, VKP

nvestigation: KKHY, SB, GB, MSR, RA, JG, MCP, SJ, AD, MJW, YE, ZA-M, KH, SAS, SWM, AS, AD, ZH, JS, YC, ABE, KHC, SY, P-LK

Visualization: KKHY, SB, GB, RA, JG, MCP, SJ, YE, ZA-M, SWM, AD

Funding acquisition: The GBM TeamLab

Project administration: KKHY, NYRA, EAC, VG, BPK, LN, DN

Supervision: KKHY, NYRA, FMW, VT, PS, KLL, EAC

Writing (draft): KKHY, SB, GB, RA, JG, MCP, SJ, MJW, KLL

Writing (review & editing): KKHY, NYRA, FMW, EAC, CB, DR, CWB, SJ, MH

All authors meet the journal authorship criteria

## Data and Materials Availability

All requests for data should be made to the corresponding authors following verification of any intellectual property or confidentiality obligations.

The mass spectrometry proteomics data have been deposited to the ProteomeXchange Consortium via the PRIDE partner repository with the dataset identifier PXD046857 and 10.6019/PXD046857. Data are available via ProteomeXchange.

## ^§^ The GBM TeamLab

Jennifer Wiley^1^, Kathryn Partridge^2^, Vasilena Gocheva^2^, Ugonma N. Chukwueke^1,3^, Franziska Michor^4^, Shahiba Ogilvie^5^, Marco Mineo^6^, Md Amin Hossain^1,7^, Jordina Rincon-Torroella^8^, Jayne Vogelzang^9^, Kimberly Lopez Vasquez^7^, Isaac H. Solomon^9^, Himanshu Soni^6^, Anna Ball^7^, Raziye Piranlioglu^6^, Daniel Triggs^7^, Alexander L. Ling^6^, Nafisa Masud^7^, Ana Montalvo Landivar^7^, Marla J. Polk^10^, Charles P. Couturier^7,11,12^, Dina Elharouni^12,13^, Jian Hu^14^, Alexandra Giantini Larsen^5^, Pratibha Sharma^15^, Christopher Douville^16^

^1^ Center for Neuro-oncology, Dana-Farber Cancer Institute, Boston, MA 02115, USA

^2^ Break *Through* Cancer, Cambridge, MA 02142, USA

^3^ Department of Medical Oncology, Dana-Farber Cancer Institute, Boston, MA 02215, USA

^4^ Department of Data Science, Dana-Farber Cancer Institute, Boston, MA; Department of Biostatistics, Harvard T.H. Chan School of Public Health, Boston, MA; Department of Stem Cell and Regenerative Biology, Harvard University, Cambridge, MA USA

^5^ Department of Neurosurgery, Memorial Sloan Kettering Cancer Center, New York, NY 10065, USA

^6^ Harvey W. Cushing Neuro-oncology Laboratories, Department of Neurosurgery, Harvard Medical School and Brigham and Women’s Hospital, Boston, MA 02115, USA

^7^ Department of Neurosurgery, Brigham and Women’s Hospital, Boston, MA 02115, USA

^8^ Department of Neurosurgery, Johns Hopkins University School of Medicine, Baltimore, MD 21287, USA

^9^ Department of Pathology, Dana-Farber Cancer Institute, Boston, MA 02215, USA

^9^ Department of Pathology, Brigham and Women’s Hospital, Boston, MA 02215, USA

^10^ Department of Immunology, University of Texas MD Anderson Cancer Center, Houston, TX 77030, USA

^11^ Institute of Medical Engineering and Sciences and Koch Institute for Integrative Cancer Research, Massachusetts Institute of Technology, Cambridge, MA; Department of Cancer Biology, Dana-Farber Cancer Institute, Boston, MA 02215 USA

^12^ Broad Institute of MIT and Harvard, Cambridge, MA, 02142

^13^ Department of Oncologic Pathology, Dana-Farber Cancer Institute, Harvard Medical School, Boston, MA, 02215 USA

^14^ Department of Cancer Biology, University of Texas MD Anderson Cancer Center, Houston, TX 77030, USA

^15^ Department of Neuro-Oncology, The University of Texas MD Anderson Cancer Center, Houston, Texas, 77054, USA

^16^ Department of Oncology, Johns Hopkins University School of Medicine, Baltimore, MD 21287, USA

## Supplementary Information

<u>Supplementary Table 1</u>

Basic patient demographic details

<u>Supplementary Table 2</u>

Dissociation results

<u>Supplementary Table 3</u>

List of antibodies for spatial proteomics

<u>Supplementary Table 4</u>

List of antibodies for CyCIF

<u>Supplementary Data File 1</u>

Immunopeptidomics

<u>Supplementary Data File 2</u>

Proteomics and phosphoproteomics

<u>Supplementary Data File 3</u>

Gene Expression and Metabolite Enrichment

<u>Supplementary Appendix 1</u>

Additional Author Affiliations for GBM TeamLab

<u>Supplementary Figure 1</u>

**TCR clonal analysis from stereotactic biopsy**

The GBM immune environment is typically a T-cell sparse environment, and measurement of the diversity and abundance of T-cell receptor (TCR) repertoire within the tumor is an important baseline measure before, during and after any immunotherapeutic intervention. We therefore sought to measure TCR clonal diversity and abundance at the bulk level from a single stereotactic biopsy specimen from a diagnostic FFPE sample from the same patient. Analysis showed approximately 500 unique CDR3 clones with approximately 1000 total TCRs. The majority of TCR clones are of size 1, and the relationship between the rank and clonal size obeys a power law, similar to what is seen in other TCR studies (*92*).

**Supplemental Fig 1.**
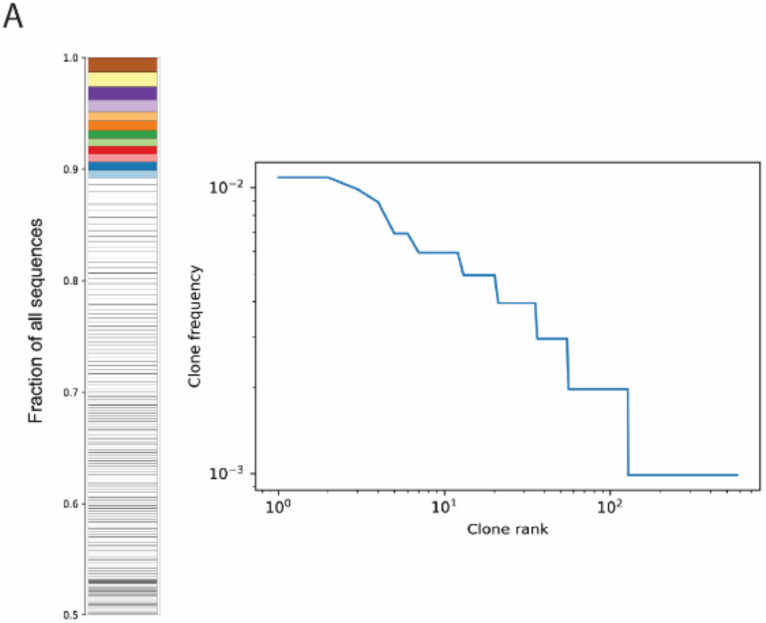
Bulk TCR sequencing data from needle core biopsy: Left) Barplot demonstrating each clone as a stacked bar. The majority of clones are size 1 clones. Right) Rank-Frequency plot showing all clones by rank order on the x-axis and fractional size on the y-axis on a log-log plot.

**Supplementary Fig 2:**
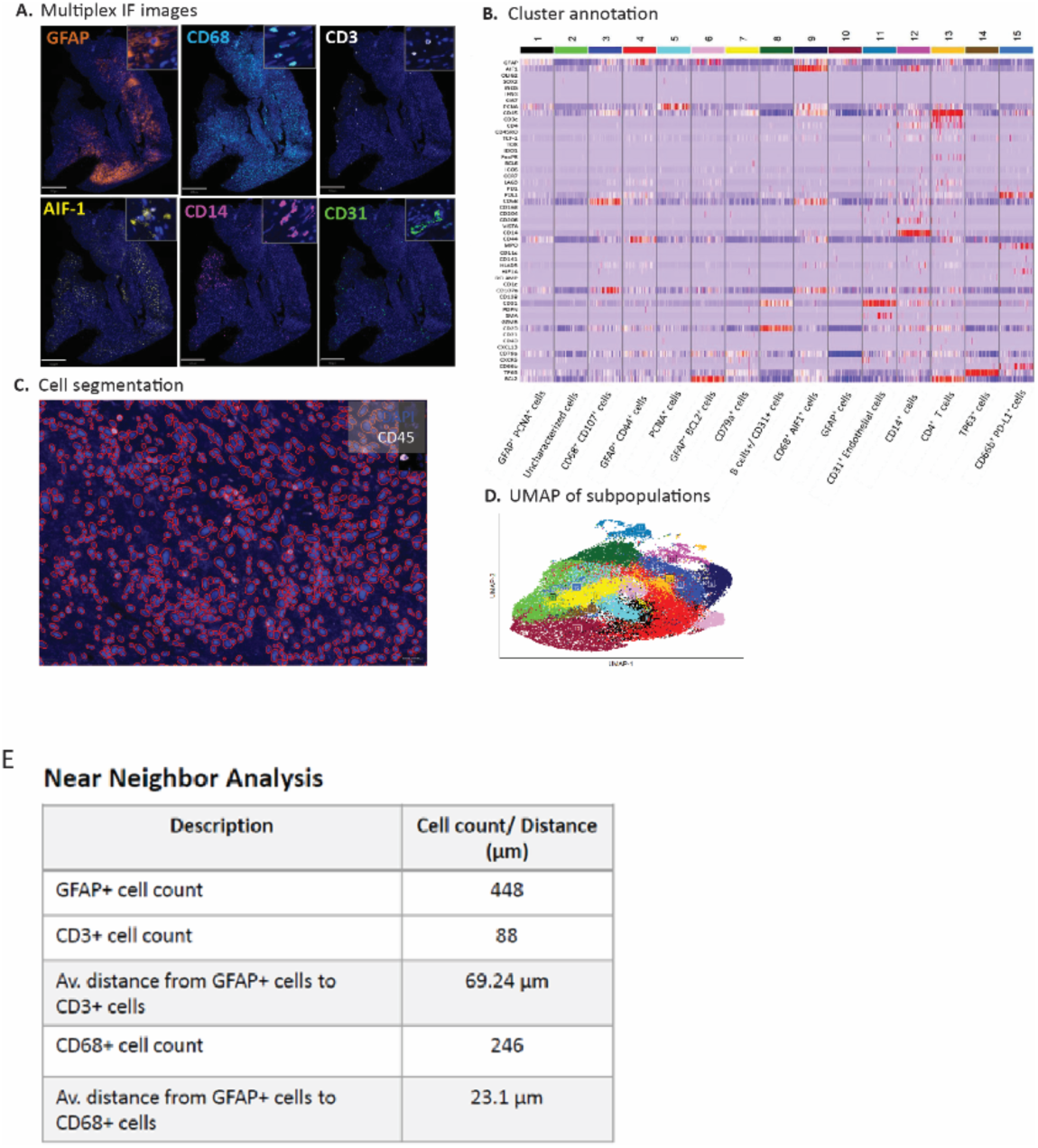
Additional CODEX analysis. A) Representative Multiplex IF images. B) Cell type clustering by CODEX markers. C) Representative cellular segmentation anlaysis. D) UMAP of cellular subpopulations as detected by CODEX. E) Cellular Neighborhood analysis metrics.

**Supplementary Table 1.**
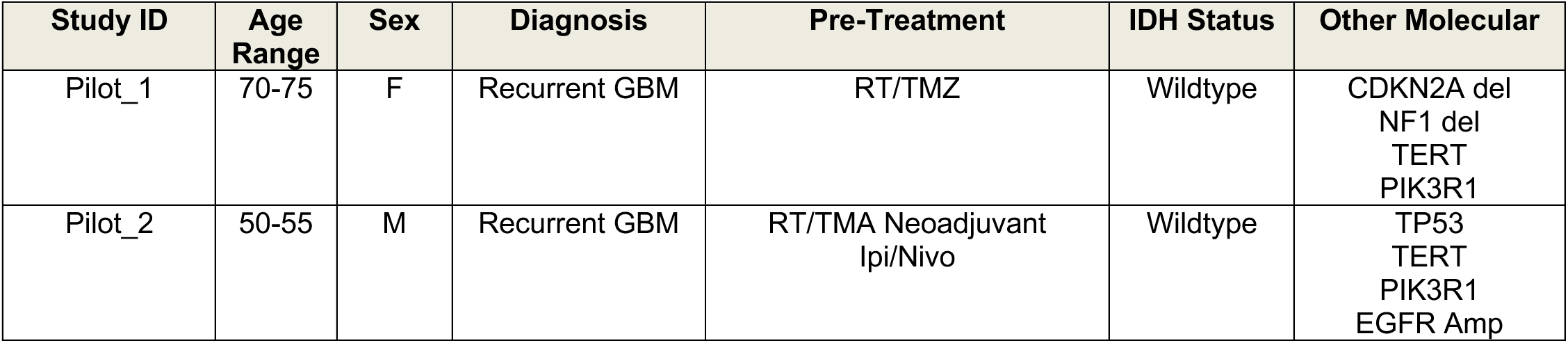
Patient Characteristics.

**Supplementary Table 2.**
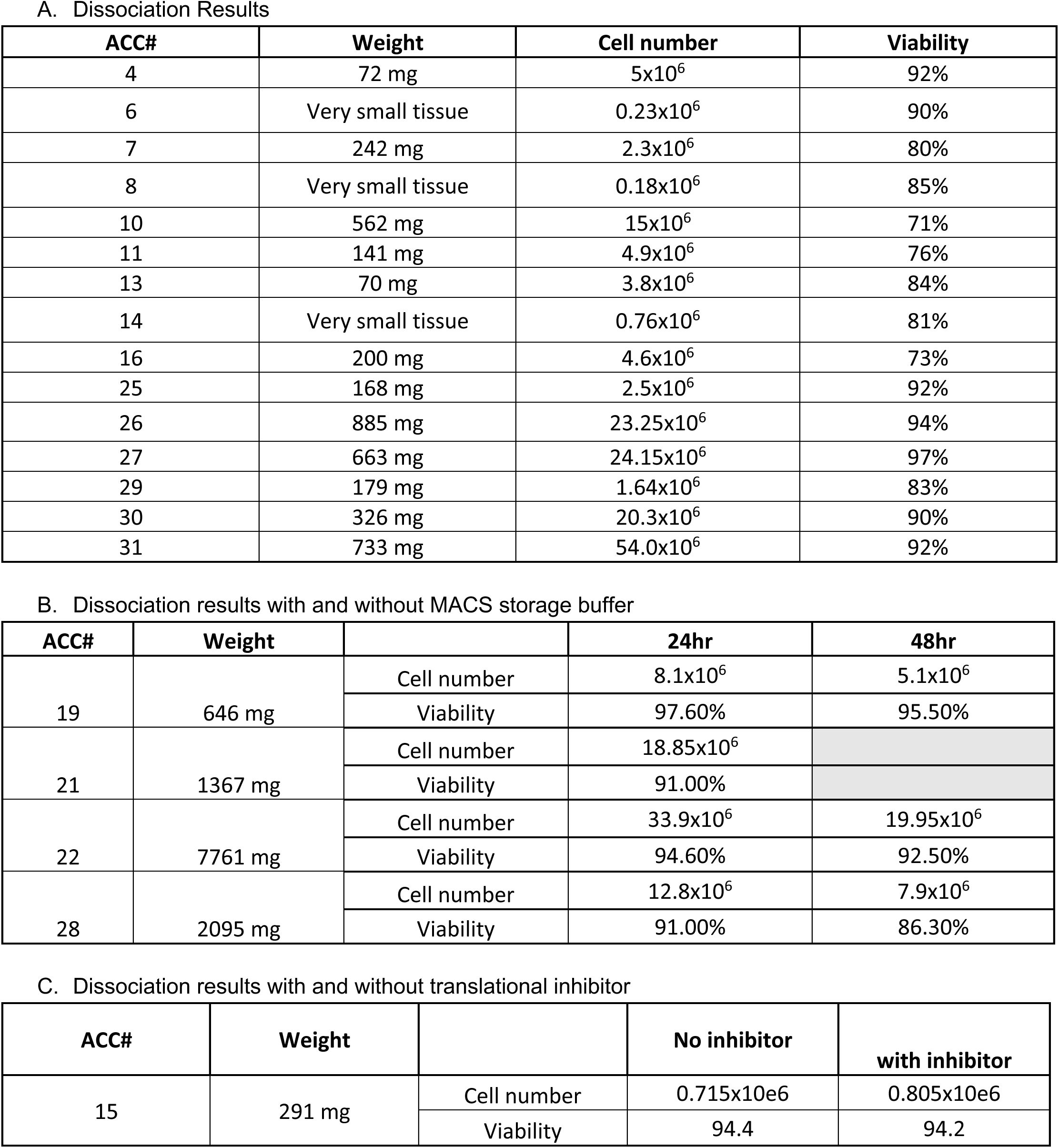

**Supplementary Table 3.**
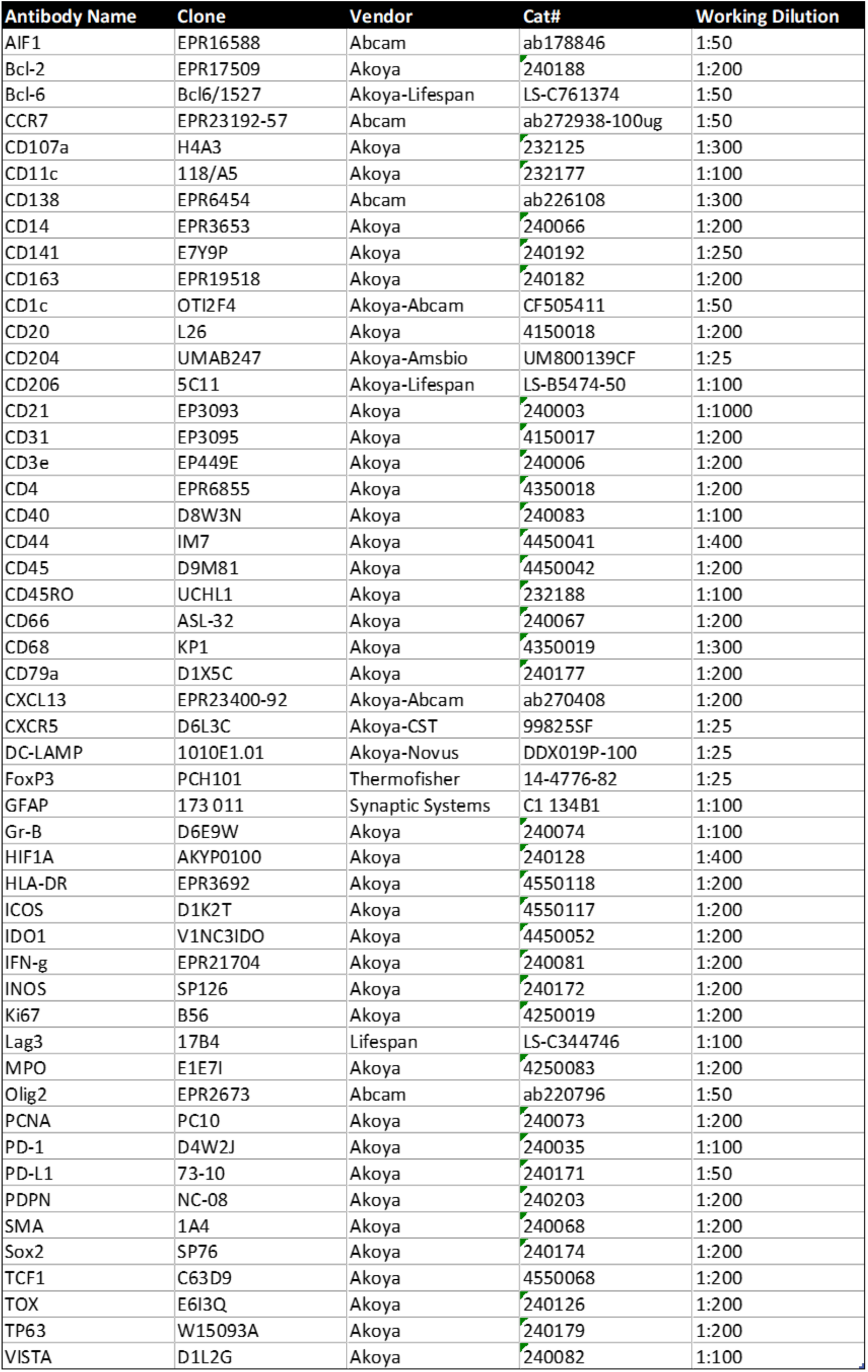
CODEX Antibodies.

**Supplementary Table 4.**
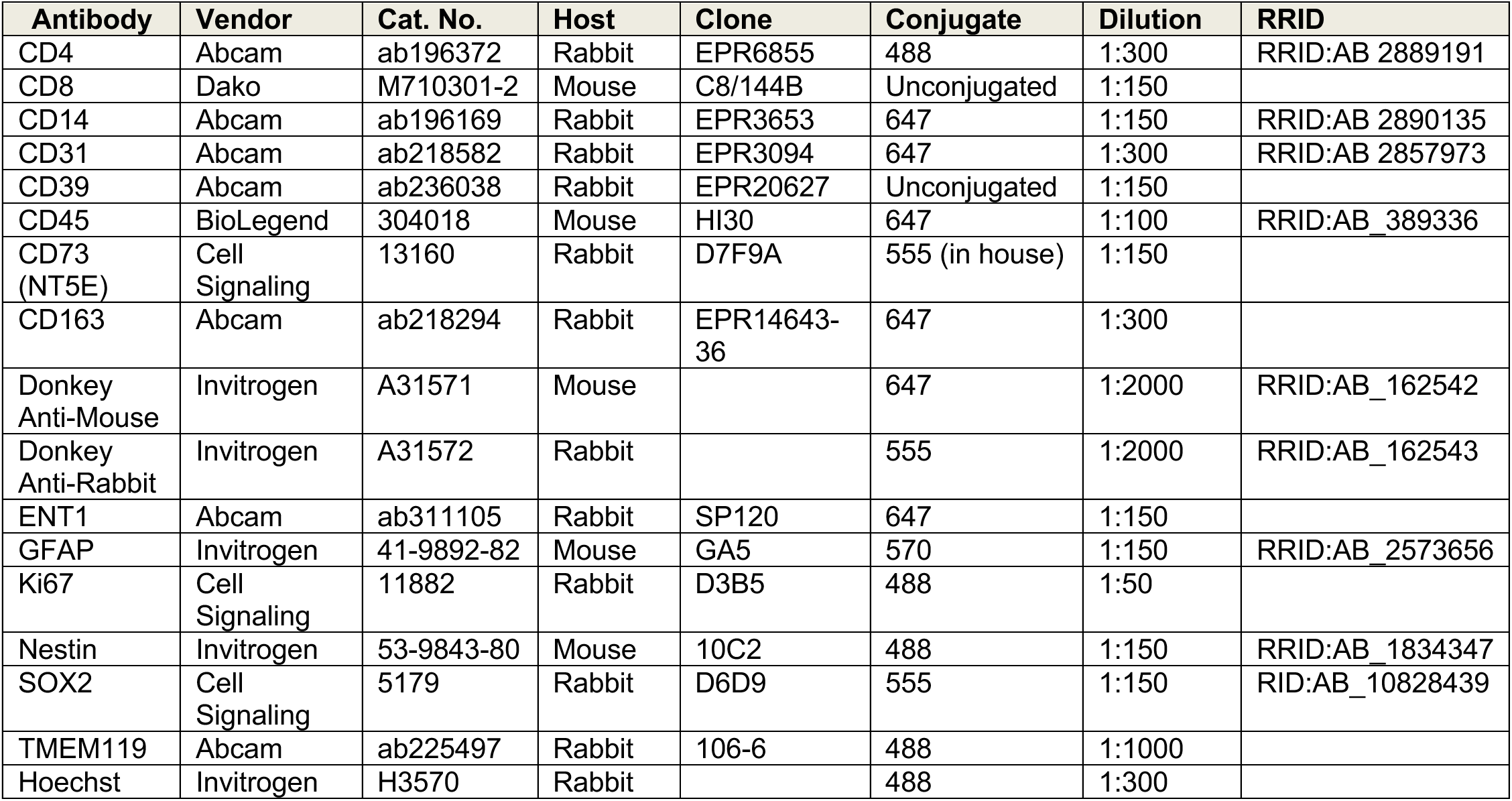
List of Antibodies for CyCIF.

