## Supplemental Information for "Investigative needle core biopsies for multi-omics in Glioblastoma"

Supplementary Figure 1

**TCR clonal analysis from stereotactic biopsy**

The GBM immune environment is typically a T-cell sparse environment, and measurement of the diversity and abundance of T-cell receptor (TCR) repertoire within the tumor is an important baseline measure before, during and after any immunotherapeutic intervention. We therefore sought to measure TCR clonal diversity and abundance at the bulk level from a single stereotactic biopsy specimen from a diagnostic FFPE sample from the same patient. Analysis showed approximately 500 unique CDR3 clones with approximately 1000 total TCRs. The majority of TCR clones are of size 1, and the relationship between the rank and clonal size obeys a power law, similar to what is seen in other TCR studies *(92)*.

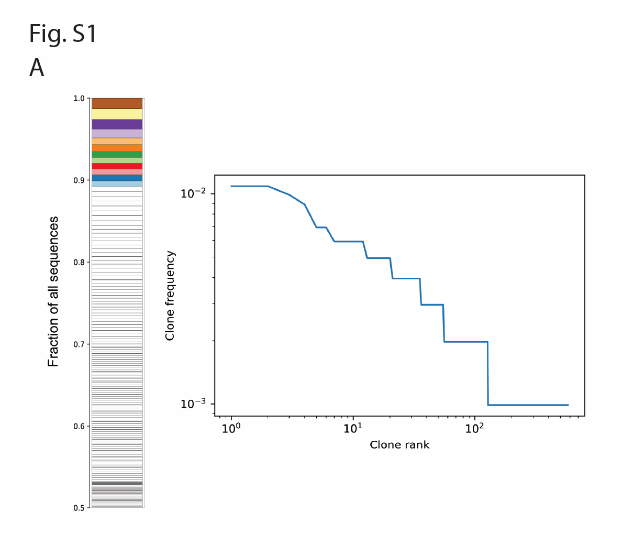

*Supplemental Fig 1. Bulk TCR sequencing data from needle core biopsy: Left) Barplot demonstrating each clone as a stacked bar. The majority of clones are size 1 clones. Right) Rank-Frequency plot showing all clones by rank order on the x-axis and fractional size on the y-axis on a log-log plot.*

Supplementary Figure 2

**CODEX analysis on patient 1 sample**

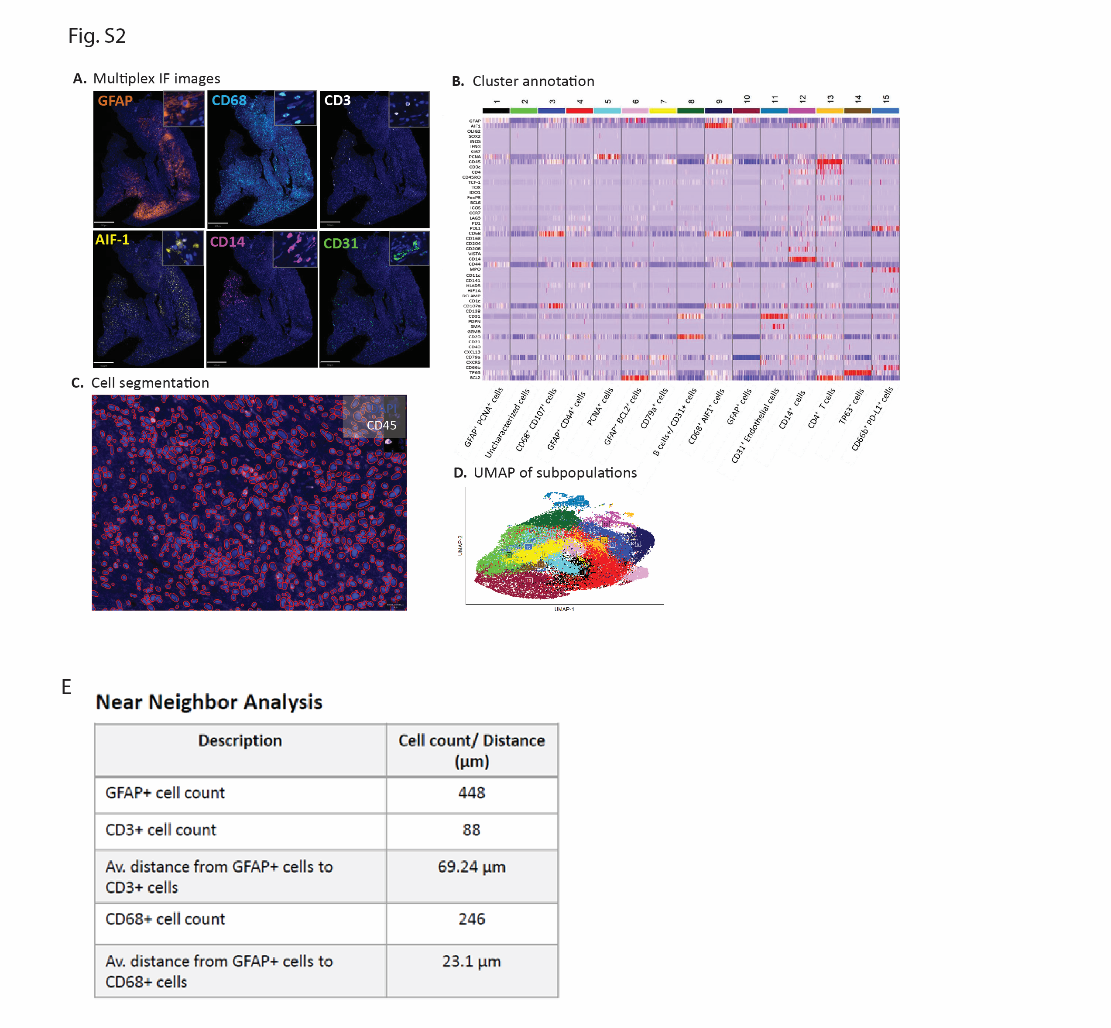

*Supplementary Fig 2: Additional CODEX analysis A) Representative Multiplex IF images. B) Cell type clustering by CODEX markers. C) Representative cellular segmentation anlaysis. D) UMAP of cellular subpopulations as detected by CODEX. E) Cellular Neighborhood analysis metrics.*

**Supplementary Table 1**

Patient Characteristics

| **Study ID** | **Age Range** | **Sex** | **Diagnosis** | **Pre-Treatment** | **IDH Status** | **Other Molecular** |
| --- | --- | --- | --- | --- | --- | --- |
| Pilot_1 | 70-75 | F | Recurrent GBM | RT/TMZ | Wildtype | CDKN2A del  NF1 del  TERT  PIK3R1 |
| Pilot_2 | 50-55 | M | Recurrent GBM | RT/TMA Neoadjuvant Ipi/Nivo | Wildtype | TP53  TERT  PIK3R1  EGFR Amp |

**Supplementary Table 2**

A. Dissociation Results

| **ACC#** | **Weight** | **Cell number** | **Viability** |
| --- | --- | --- | --- |
| 4 | 72 mg | 5x10^6^ | 92% |
| 6 | Very small tissue | 0.23x10^6^ | 90% |
| 7 | 242 mg | 2.3x10^6^ | 80% |
| 8 | Very small tissue | 0.18x10^6^ | 85% |
| 10 | 562 mg | 15x10^6^ | 71% |
| 11 | 141 mg | 4.9x10^6^ | 76% |
| 13 | 70 mg | 3.8x10^6^ | 84% |
| 14 | Very small tissue | 0.76x10^6^ | 81% |
| 16 | 200 mg | 4.6x10^6^ | 73% |
| 25 | 168 mg | 2.5x10^6^ | 92% |
| 26 | 885 mg | 23.25x10^6^ | 94% |
| 27 | 663 mg | 24.15x10^6^ | 97% |
| 29 | 179 mg | 1.64x10^6^ | 83% |
| 30 | 326 mg | 20.3x10^6^ | 90% |
| 31 | 733 mg | 54.0x10^6^ | 92% |

B. Dissociation results with and without MACS storage buffer

| **ACC#** | **Weight** |  | **24hr** | **48hr** |
| --- | --- | --- | --- | --- |
| 19 | 646 mg | Cell number | 8.1x10^6^ | 5.1x10^6^ |
|  |  | Viability | 97.60% | 95.50% |
| 21 | 1367 mg | Cell number | 18.85x10^6^ |  |
|  |  | Viability | 91.00% |  |
| 22 | 7761 mg | Cell number | 33.9x10^6^ | 19.95x10^6^ |
|  |  | Viability | 94.60% | 92.50% |
| 28 | 2095 mg | Cell number | 12.8x10^6^ | 7.9x10^6^ |
|  |  | Viability | 91.00% | 86.30% |

C. Dissociation results with and without translational inhibitor

| **ACC#** | **Weight** |  | **No inhibitor** | **with inhibitor** |
| --- | --- | --- | --- | --- |
| 15 | 291 mg | Cell number | 0.715x10e6 | 0.805x10e6 |
|  |  | Viability | 94.4 | 94.2 |

**Supplementary Table 3**

CODEX Antibodies

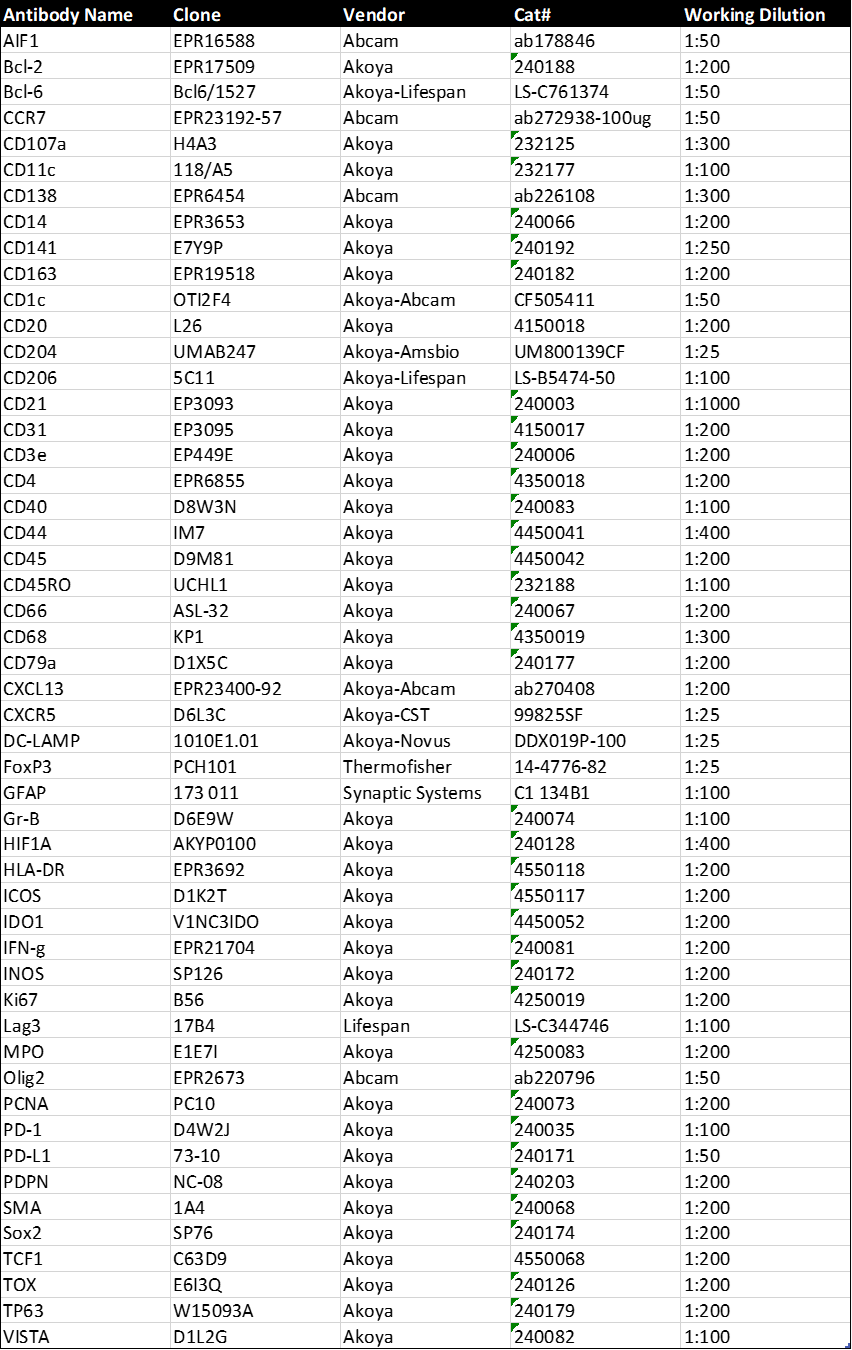

**Supplementary Table 4**

List of Antibodies for CyCIF

| **Antibody** | **Vendor** | **Cat. No.** | **Host** | **Clone** | **Conjugate** | **Dilution** | **RRID** |
| --- | --- | --- | --- | --- | --- | --- | --- |
| CD4 | Abcam | ab196372 | Rabbit | EPR6855 | 488 | 1:300 | RRID:AB 2889191 |
| CD8 | Dako | M710301-2 | Mouse | C8/144B | Unconjugated | 1:150 |  |
| CD14 | Abcam | ab196169 | Rabbit | EPR3653 | 647 | 1:150 | RRID:AB 2890135 |
| CD31 | Abcam | ab218582 | Rabbit | EPR3094 | 647 | 1:300 | RRID:AB 2857973 |
| CD39 | Abcam | ab236038 | Rabbit | EPR20627 | Unconjugated | 1:150 |  |
| CD45 | BioLegend | 304018 | Mouse | HI30 | 647 | 1:100 | RRID:AB_389336 |
| CD73 (NT5E) | Cell Signaling | 13160 | Rabbit | D7F9A | 555 (in house) | 1:150 |  |
| CD163 | Abcam | ab218294 | Rabbit | EPR14643-36 | 647 | 1:300 |  |
| Donkey Anti-Mouse | Invitrogen | A31571 | Mouse |  | 647 | 1:2000 | RRID:AB_162542 |
| Donkey Anti-Rabbit | Invitrogen | A31572 | Rabbit |  | 555 | 1:2000 | RRID:AB_162543 |
| ENT1 | Abcam | ab311105 | Rabbit | SP120 | 647 | 1:150 |  |
| GFAP | Invitrogen | 41-9892-82 | Mouse | GA5 | 570 | 1:150 | RRID:AB_2573656 |
| Ki67 | Cell Signaling | 11882 | Rabbit | D3B5 | 488 | 1:50 |  |
| Nestin | Invitrogen | 53-9843-80 | Mouse | 10C2 | 488 | 1:150 | RRID:AB_1834347 |
| SOX2 | Cell Signaling | 5179 | Rabbit | D6D9 | 555 | 1:150 | RID:AB_10828439 |
| TMEM119 | Abcam | ab225497 | Rabbit | 106-6 | 488 | 1:1000 |  |
| Hoechst | Invitrogen | H3570 | Rabbit |  | 488 | 1:300 |  |
